# Single-cell dissection of schizophrenia reveals neurodevelopmental-synaptic axis and transcriptional resilience

**DOI:** 10.1101/2020.11.06.20225342

**Authors:** W. Brad Ruzicka, Shahin Mohammadi, Jose Davila-Velderrain, Sivan Subburaju, Daniel Reed Tso, Makayla Hourihan, Manolis Kellis

**Affiliations:** Laboratory for Epigenomics in Human Psychopathology, McLean Hospital, Belmont, MA, USA; Harvard Medical School, Boston, MA, USA; Broad Institute of MIT and Harvard, Cambridge, MA, USA; MIT Computer Science and Artificial Intelligence Laboratory, Cambridge, MA, USA

**Author notes:** These authors contributed equally: W. Brad Ruzicka, Shahin Mohammadi.

## Abstract

Schizophrenia is a devastating mental disorder with a high societal burden, complex pathophysiology, and diverse genetic and environmental risk factors. Its complexity, polygenicity, and small-effect-size and cell-type-specific contributors have hindered mechanistic elucidation and the search for new therapeutics. Here, we present the first single-cell dissection of schizophrenia, across 500,000+ cells from 48 postmortem human prefrontal cortex samples, including 24 schizophrenia cases and 24 controls. We annotate 20 cell types/states, providing a high-resolution atlas of schizophrenia-altered genes and pathways in each. We find neurons are the most affected cell type, with deep-layer cortico-cortical projection neurons and parvalbumin-expressing inhibitory neurons showing significant transcriptional changes converging on genetically-implicated regions. We discover a novel excitatory-neuron cell-state indicative of transcriptional resilience and enriched in schizophrenia subjects with less-perturbed transcriptional signatures. We identify key trans-acting factors as candidate drivers of observed transcriptional perturbations, including MEF2C, TCF4, SOX5, and SATB2, and map their binding patterns in postmortem human neurons. These factors regulate distinct gene sets underlying fetal neurodevelopment and adult synaptic function, bridging two leading models of schizophrenia pathogenesis. Our results provide the most detailed map to date for mechanistic understanding and therapeutic development in neuropsychiatric disorders.

## Introduction

Schizophrenia afflicts young adults just as they approach their full potential, manifests as a combination of psychosis, social withdrawal, and cognitive dysfunction, and often leads to a lifetime of profound and chronic disability^1^. Schizophrenia pathogenesis is thought to begin during neurodevelopment, yet symptoms do not emerge until many years later in early adulthood^2^. The complex etiology of schizophrenia is driven by both genetic and environmental factors affecting a wide range of brain-related processes, including neurodevelopment^3,4^, cognition^5,6^, synaptic function^7,8^, neuronal excitability^9,10^, and neuronal connectivity^11,12^.

Decades-long efforts to decipher schizophrenia genetics have yielded 145+ robustly-associated genetic loci^13,14^, but their target genes and cell-types of action remain largely uncharacterized, hindering the search for mechanistic insights and therapeutics development. Cell-type-specific profiles of reference (non-schizophrenia) brain-sample transcriptomes have shown that genes near schizophrenia risk loci are expressed in pyramidal neurons and specific subsets of inhibitory neurons^15^, but do not reveal whether schizophrenia-locus-proximal genes are indeed differentially-expressed in schizophrenia, or whether increased or decreased expression is risk-associated or protective. Conversely, schizophrenia case-control gene expression analyses using homogenized cortical tissue have been carried out at the bulk level^16–19^, revealing several differentially-expressed genes in schizophrenia. However, such bulk-level studies average gene expression across millions of cells, merging together diverse and inconsistent cell types, and thus miss genes that are differentially-expressed only in lower-abundance cell types and genes with opposite changes in different cell populations, and can also result in false positives stemming from cell-type-composition changes between samples. Emerging technologies for single-cell transcriptomics^20,21^ achieve both cell-type-specificity and reveal disease-associated changes, as demonstrated for Alzheimer’s Disease^22^, Autism Spectrum Disorder^23^, Major Depressive Disorder^24^, and Multiple Sclerosis^25^, but these have not been applied to schizophrenia to date.

Here, we present the first transcriptomic atlas of schizophrenia at single-cell resolution, profiling >500,000 cells from postmortem human prefrontal cortex from 24 schizophrenia, and 24 age-matched control individuals. We annotate 18 distinct cell types, including seven excitatory and six inhibitory neuronal subpopulations and five non-neuronal cell types. We also identify a new excitatory neuron cell-state, Ex-SZTR, which is enriched for differentially-expressed genes and significantly more prevalent in schizophrenia than in control individuals, but surprisingly preferentially found in schizophrenia individuals with non-schizophrenia transcriptional signatures across all other cell types, indicating transcriptional resilience. Schizophrenia-associated transcriptional perturbations enrich in highly-specific cellular populations, including deep-layer cortico-cortical projection neurons and PV-expressing inhibitory neurons, highlighting the importance of cell-type-specific assessments. These changes preferentially perturb postsynaptic organization, synaptic plasticity, and neurodevelopmental pathways, providing mechanistic insights into functional pathways that underlie schizophrenia perturbations. Genes showing differential expression in schizophrenia are significantly enriched in the proximity of GWAS-associated genes^14^ and linked to schizophrenia-associated genetic variants via enhancer-promoter loops in adult dorsolateral prefrontal cortex^26^, providing candidate target genes, the directionality of effect, and cell-type-of-action for 68 of 145 schizophrenia loci. Searching for common upstream regulators for differentially-expressed genes, we find that schizophrenia-associated transcriptional perturbations converge on a small number of transcription factors, which are themselves encoded in schizophrenia-associated genetic loci, including MEF2C, SATB2, SOX5, and TCF4. These factors act in both early fetal brain development and in adult brain synaptic processes, thus linking two leading models of schizophrenia pathogenesis. Our results provide both a unique resource for understanding the molecular biology of schizophrenia at single-cell resolution, and numerous new insights for understanding candidate cell-type-specific driver genes, biological pathways, master regulators, and resilience mechanisms, and for prioritizing target genes for therapeutic development against this devastating disorder.

### Multiplexed single-cell profiling of schizophrenia

To investigate schizophrenia-associated cell-type-specific transcriptional disruption within the complex cytoarchitecture of the human brain, we used single-nucleus RNA sequencing (snRNA-seq) to profile postmortem tissue samples from the prefrontal cortex (Brodmann Area 10) (**Fig. 1a**). We curated a cohort of 24 schizophrenia and 24 control subjects, balanced for gender (12 male and 12 female subjects per group), age (ranging from 22 to 94 years, average 63.5 years), and postmortem interval (ranging from 6.9 to 26.3 hours, average 16.8 hours) (**Supplementary Table 1**). We reviewed medical records and incorporated psychiatric medication exposures into our analysis to control for potential confounding effects of pharmacotherapy. We multiplexed samples, pooling three schizophrenia and three control samples in each of eight batches, using the Multiplexing Using Lipid Tagged Indices (MULTI-Seq)^27^. Compared to standard snRNA-seq protocols, sample multiplexing allows us to capture more cells from each individual while reducing batch effects (by profiling both schizophrenia and control samples in the same sequencing library), reducing the rate of undetected doublets (by using sample hashtags to remove cross-individual doublets), and lowering sample preparation costs (by assessing more than one sample per 10x kit channel). After doublet removal, we obtained 560,020 single-nucleus transcriptomes, including 266,431 cells from schizophrenia and 293,589 from control individuals, profiled at an average depth of 12k cells per sample and 35k reads per cell (**Supplementary Table 2**). We next removed genes expressed in less than ten cells in each batch, and cells with fewer than 500 identified genes or more than 10% of unique molecular identifiers (UMIs) in mitochondrial genes, resulting in 17,460 genes detected in 506,378 cells.

**Figure 1.**
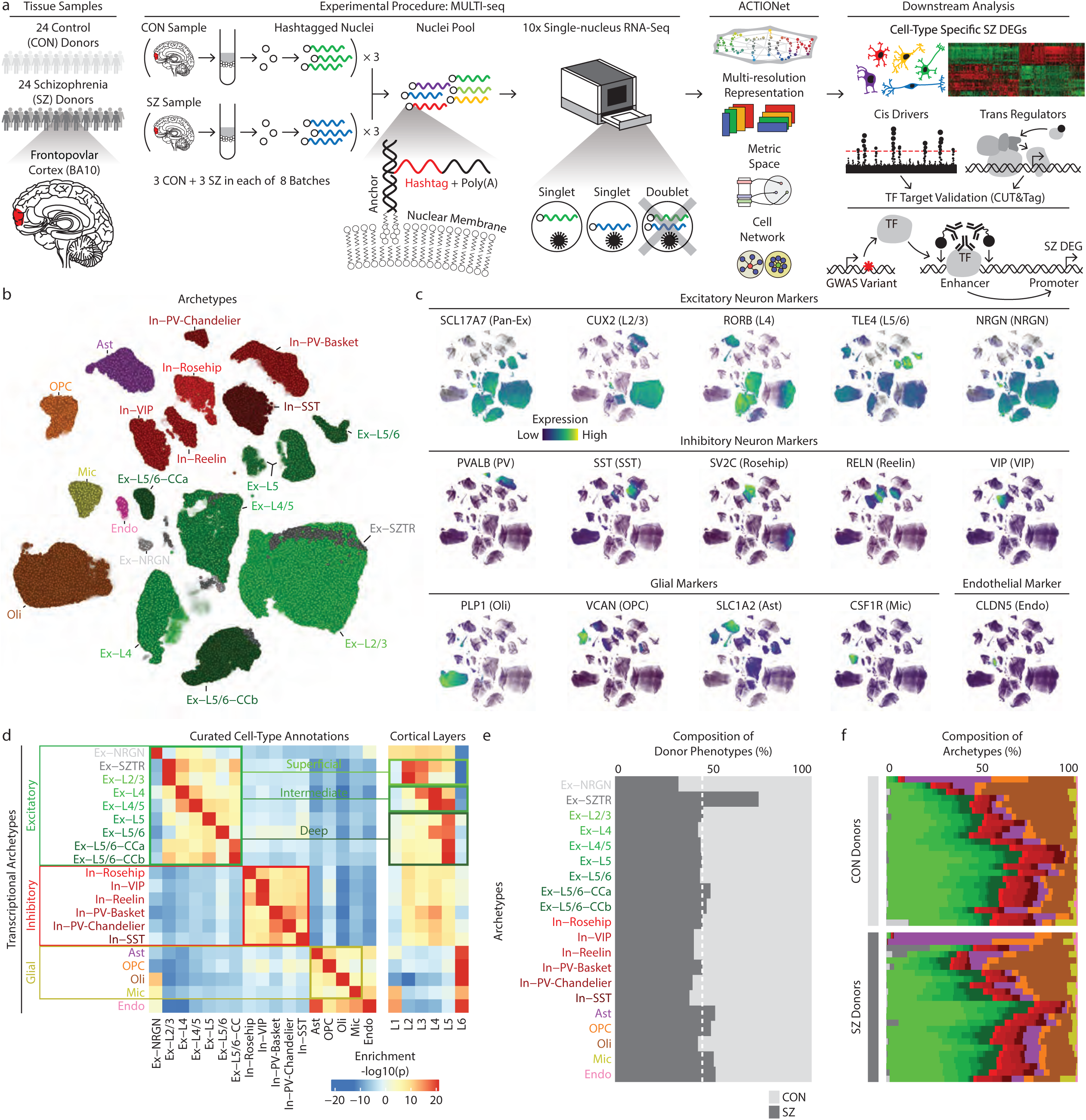
Multiresolution dissection of cellular subpopulations. **a**. Overview of study design and data analysis strategies. TF - transcription factor. CUT&Tag - Cleavage Under Targets and Tagmentation. **b**. ACTIONet plot of putative cell types/states. Green and red clusters represent excitatory and inhibitory subtypes of neurons respectively, with darker shades indicating an association with deeper cortical layers. **c**. Projection of known marker genes verifies cell type annotations and cortical layer associations^82^. **d**. Annotation of transcriptional archetypes using curated markers from previous studies. **e**. The percentage of cells within each cell-type/state contributed by SZ and CON subjects. **f**. Cell-type/state decomposition of individual samples. Colors are consistent with those used in panel b.

### Multiresolution dissection of cellular subpopulations

We used our recently developed multiresolution cell-state discovery platform, ACTIONet^28^, to identify both discrete and continuous cell states (“archetypes”) using a coupled archetypal/network analysis. We first applied ACTIONet to reconstruct the geometry of the cell-state landscape and used it to detect and remove outlier cells that fall on the periphery of the constructed cell-cell similarity graph, keeping only the 386,065 highest-quality cells with reproducible expression patterns. In the second round, we used the ACTIONet on the filtered cells and identified 20 cell states in the combined population of schizophrenia and control subjects, nearly all of which form dense clusters in the cell-cell similarity network (**Fig. 1b, Extended Data Fig. 1a**), with two notable exceptions that we discuss below. Our 20 cell states captured all major cell types of the human prefrontal cortex, including subtypes of excitatory and inhibitory neurons and glia (**Fig. 1b, d**). We verified these annotations based on known marker gene expression patterns, by projecting individual marker genes on the cell similarity network (**Fig. 1c**). *De novo* markers for these cell types are consistent with our previous studies of the human prefrontal cortex^28^ (**Supplementary Table 3**). Neuronal subtypes, particularly excitatory neurons, showed higher numbers of expressed genes and identified UMIs (**Extended Data Fig. 2a, b**).

We also captured all major subtypes of GABAergic inhibitory neurons, including calcium-binding protein parvalbumin (PV) expressing neurons, neuropeptide somatostatin (SST) neurons, and ionotropic serotonin receptor 5HT3a (5HT3aR) neurons. Within these groups, we detected two PV-expressing subtypes of inhibitory neurons (Basket and Chandelier), and two 5HT3aR-expressing subtypes (VIP+ and Reelin+), and the recently-described Rosehip population^29^. Cell groupings observed in the ACTIONet plot are consistent with the developmental origin of cardinal interneuron subtypes (medial versus caudal ganglionic eminence), demonstrating their shared transcriptional signatures^30^.

Excitatory neurons exhibit a strong layer-specific pattern. We observed a consistent association of superficial-layer excitatory neurons (layers II/II) with marker genes *CUX2* and *CBLN2*. However, intermediate and deep-layer excitatory neurons are more intermixed, with layer IV/V neurons enriched for *RORB, FOXP2*, and *RXFP1*, while deep layer neurons are marked with *TLE4, SEMA3E*, and *HTR2C* genes. Within the deep cortical layers V and VI, we observe three distinct populations of excitatory neurons: cortico-fugal projection neurons (Ex-L5/6) expressing *FEZF2*, and two distinct populations of cortico-cortical projection neurons (Ex-L5/6-CCa, Ex-L5/6-CCb).

To further investigate differences between Ex-L5/6-CCa and Ex-L5/6-CCb, we focused on cells from control individuals associated with these cell-types and performed differential expression analysis (Wilcoxon’s rank-sum test, **Supplementary Table 4**). We found that Ex-L5/6-CCa is enriched for genes involved in the dopamine receptor signaling pathway, whereas Ex-L5/6-CCb is enriched for many key genes in glutamate signaling. These distinct expression profiles point to physiological differences between two deep layer neuronal populations likely related to specifics of their afferent and efferent connectivity with distinct cortical and subcortical regions.

In addition to major cell types and their corresponding subtypes, we found two transcriptional archetypes that capture continuous cell states shared across multiple subtypes of excitatory neurons. The first cross-cutting cell state (Ex-NRGN) captures both a localized cell-type characterized by the expression of the *NRGN* gene described in prior snRNA-seq studies of postmortem human brain^23^, as well as a transcriptional signature distributed among cells of different subtypes. The Ex-NRGN cell-state does not show enrichment for layer-specific markers. In addition to *NRGN*, this population is marked by expression of the Brain Expressed and X-Linked gene family members (*BEX1* and *BEX3*), calcium-binding enzymatic cofactor Calmodulin 3 (*CALM3*), and *YWHAH*, which encodes a 14-3-3 signal transduction protein implicated in the conversion to the psychosis of at-risk individuals^31^. Cells associated with the Ex-NRGN cell state highly express mitochondrial genes (**Extended Data Fig. 2c**), an observation reproducible in an independent study^23^ (**Extended Data Fig. 2d**).

The second cross-cutting cell-state (Ex-SZTR) is of particular interest, as it is preferentially associated with excitatory neurons from schizophrenia patients (**Fig. 1e**). Ex-SZTR cells are enriched in superficial layer excitatory neurons, as evidenced by the expression of layer-specific marker genes as well as the localization of these cells within the cell network (**Fig. 1b**,**d, Extended Data Fig. 1a**). The top-ranked associated genes are enriched for pathways related to synapse organization and synaptic plasticity, as well as neuronal process morphology (**Supplementary Table 5**). We name this cell state Ex-SZTR for “schizophrenia transcriptional resilience” as it is preferentially found in schizophrenia individuals whose transcriptional profiles are surprisingly non-schizophrenia-like, as we discuss later in the text.

### Cell-type-specific schizophrenia-associated transcriptional perturbations

Across all cell types/states, we identified a total of 1,637 up-regulated and 2,492 down-regulated differentially expressed genes (DEGs) in schizophrenia (3,742 distinct schizophrenia-perturbed genes, as 387 genes are both up- and down-regulated in distinct cell types), with the highest number of up-regulated genes in Ex-SZTR and the highest number of downregulated genes observed in Ex-L5/6-CCb (**Fig. 2a, Supplementary Table 6**). The majority of observed perturbations occur in neuronal populations, and ∼60% of observed perturbations are downregulation events. Among inhibitory neurons, PV-expressing subtypes (both Chandelier and Basket) which have been the focus of much prior work^32^, indeed show the highest number of perturbed genes. Pairwise comparison of the schizophrenia-associated transcriptional dysregulation between different cell subpopulations reveals similarity in transcriptional changes within all neuronal subpopulations. Among neuronal cell types, we observed a particularly high concordance of dysregulated genes among superficial layer excitatory neurons (Ex-L2/3, Ex-L4, and Ex-L4/5) and Ex-L5/6-CCb (**Extended Data Fig. 3**).

**Figure 2.**
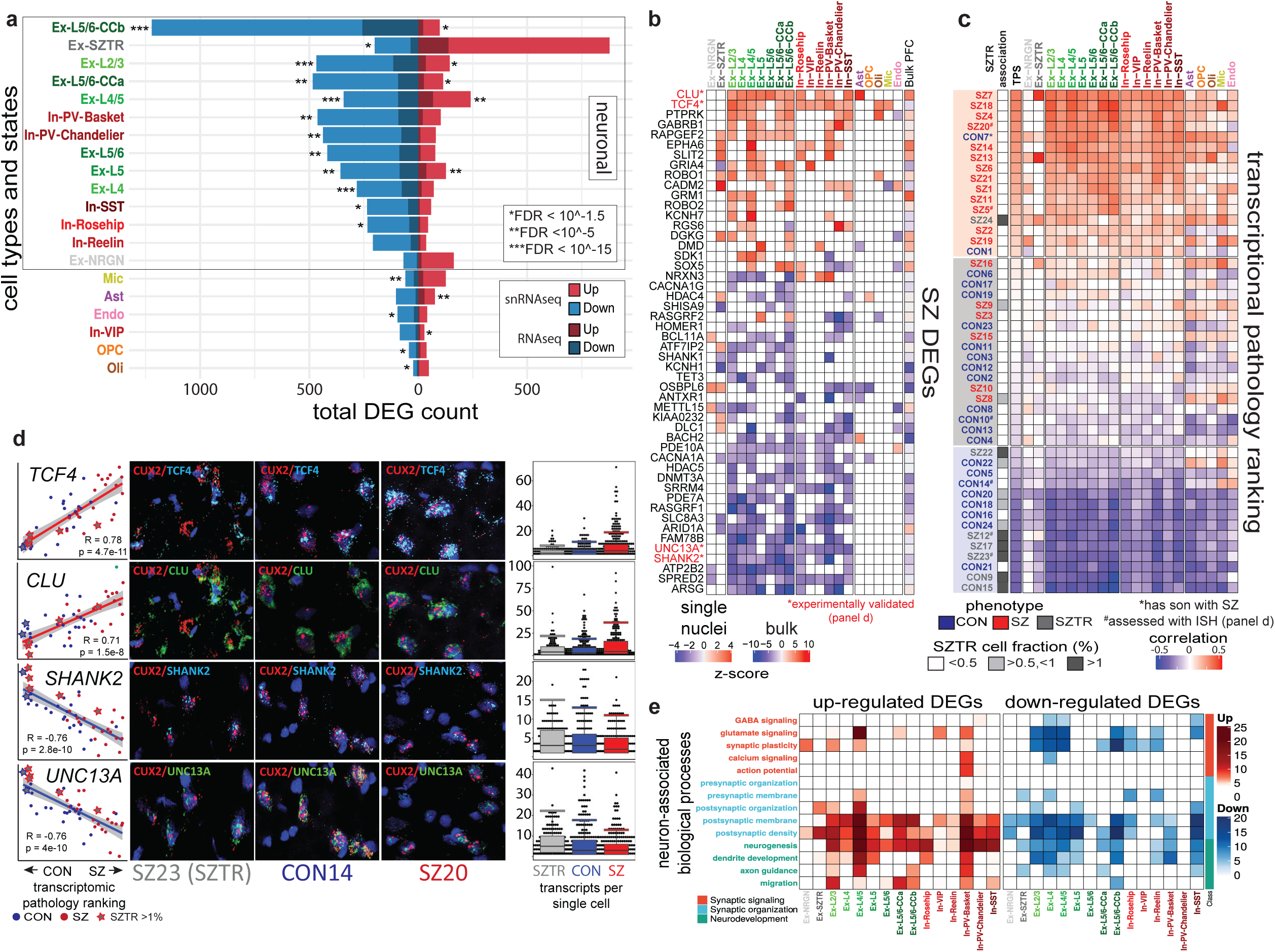
Differential gene expression analysis. **a**. Total count of up- and downregulated genes in each cell-type/state and the overlap of cell-type/state-specific SZ DEGs with DEGs observed in previous studies of bulk cortical tissue^28^.**b**. Cell-type-specific differential expression of selected top-ranked genes across all 20 cell-types/states as well as previous studies of bulk cortical tissue. Many selected genes demonstrate consistent up or downregulation across most cell populations, while others show distinct patterns across major categories (*NRXN3, RASGRF2, BACH2, DMD*), and several genes are dysregulated in opposite directions within Ex-SZTR and multiple neuronal cell-types (*GRIA4, DLC1, KIAA0232, HDAC4, ATF7IP2*). Blue indicates downregulation, and red indicates overexpression in SZ. Comparisons with nominal p>0.05 are not colored.**c**. Ranking of individuals based on an aggregate Transcriptional Pathology Score computed across all neuronal cell-types/states. Red indicates a transcriptomic signature more typical of SZ, while blue indicates a signature more typical of CON. *Individual CON7 has a first degree relative with SZ. **d** The first column depicts the relative pseudobulk expression across all neuronal cell-types of *TCF4, CLU, SHANK2*, or *UNC13A* on the y-axis plotted against the transcriptional pathology score ranking of each subject on the x-axis. Columns two through four show representative photomicrographs of fluorescent in situ hybridization for *CUX2* in red, a marker of excitatory neurons in layers II and III of the cortex, and *TCF4, CLU, SHANK2*, or *UNC13A* in blue or green. Column five depicts quantitation of the in situ hybridization signal as detected transcripts per single *CUX2* positive cell using DotDotDot^76^. Points indicate individual cell counts, boxes indicate the median and the interquartile range, and whiskers indicate the largest value within 1.5 times the interquartile range above the 75th percentile. **e**. Functional enrichment of cell-type/state-specific SZ DEGs within neuronally relevant biological processes.

To evaluate the reproducibility of our cell-type-specific dysregulated genes, we compared them to bulk-level dysregulated genes across 559 schizophrenia and 936 control homogenized prefrontal cortex samples^19^. We observed a significant overlap between the 2,450 upregulated (*p-*value<10^−8^, Fisher’s exact test) and 2,371 downregulated (*p*-value<10^−29^) bulk-level genes and the upregulated and downregulated gene sets identified here, with downregulated genes in excitatory neurons having the strongest association (**Fig. 2a**). However, among our cell-type-specific DEGs we identify novel genes not detected in prior tissue-level observations, including genes in the Ex-SZTR cell state and subtypes of inhibitory neurons, highlighting the importance of single-cell resolution analysis.

Among our DEGs (**Fig. 2b**), the transcriptional regulator *TCF4*^*14*^, which lies in a schizophrenia-associated genetic locus, was the most widely-perturbed gene, upregulated in 14 of 20 cell types. Indeed, *TCF4* was also detected as upregulated in bulk tissue RNA-Seq^18^. *TCF4* plays critical roles in both neurodevelopment and in regulating the excitability of prefrontal cortex neurons^33^, and was previously predicted as a “master regulator” of schizophrenia gene network perturbations^34^. Additionally, the molecular chaperone *CLU*, which also lies in a schizophrenia-associated genetic locus^14^ and is most prominently expressed in astrocytes, was broadly overexpressed across astrocytes, all excitatory neuron cell types, and most inhibitory neuron cell types. Confirming our findings, *CLU* was previously recognized to be hypomethylated in schizophrenia bulk postmortem brain samples^35^, and overexpressed in laser-capture-microdissected excitatory neurons and in parvalbumin-expressing inhibitory neurons^36,37^. We also observed two gene families, neurexins (*NRXN1, NRXN2, NRXN3*) and *SHANK* genes, to be commonly dysregulated across multiple neuronal subpopulations. Neurexin genes serve as presynaptic cell-adhesion molecules and interact with neuroligins within the postsynaptic membrane to mediate multiple aspects of synapse formation, structure, and function^38^. In contrast, *SHANK* genes encode postsynaptic scaffolding proteins essential for the organization of glutamatergic synapses^39^.

### Multi-gene transcriptional pathology score

We next calculated a “transcriptional pathology score” (TPS) for each individual, based on the correlation of their expression deviation (relative to the mean of all individuals) in each cell type, with the vector of schizophrenia-associated differential expression in that cell type (**Fig. 2c, Extended Data Fig. 4, Supplementary Table 7**). As expected, schizophrenia individuals showed significantly higher transcriptional pathology scores than control individuals lying at opposite extremes of the TPS distribution, indicating agreement between our predicted dysregulation scores and known phenotypic states. Individual CON7 was an exception to this trend, showing schizophrenia-like expression profiles but lacking a schizophrenia diagnosis; however, CON7 had a first-degree relative (his son) diagnosed with schizophrenia, suggesting a higher genetic predisposition than expected possibly driven by a familial strong-effect genetic alteration. Surprisingly, we found that TPS ranking was consistent across all neuronal cell types, rather than confined to the deep layer excitatory and PV-expressing basket cell-types that showed the highest number of DEGs, indicating that global pan-transcriptome gene-expression signatures of schizophrenia are robust across all neuronal cell-types (glial cell types did not show such consistency).

Strikingly, schizophrenic individuals with an abundant subset of excitatory neurons in the Ex-SZTR cell state showed among the lowest transcriptional pathology scores, even lower than most control individuals. In fact, the two control individuals marked by the Ex-SZTR state show the two lowest schizophrenia transcriptional pathology scores. This suggests that the presence of the Ex-SZTR cell state is correlated with “transcriptional resilience”, whereby even schizophrenia cases show transcriptional profiles consistent with control individuals, and control individuals show even more non-schizophrenia-like transcriptional signatures. We do not find the Ex-SZTR cell-state to be associated with exposure to specific medications or levels of exposure to pharmacologic classes. Instead, we interpret it as a potential mechanism of transcriptional compensation or resilience to schizophrenia at the molecular level worthy of further investigation.

### Experimental validation of differentially expressed genes

We next used fluorescence *in situ* hybridization (RNAscope) to experimentally validate both the cell-type-specific expression and the differential expression of four differentially-expressed genes across six individuals. We selected two upregulated genes (*TCF4, CLU*) and two down-regulated genes (*SHANK2, UNC13A*), which showed strong and consistent changes across the majority of neuronal cell-types, and validated them in two schizophrenia, two control, and two transcriptionally-resilient schizophrenia individuals. The resulting images (**Fig. 2d, Extended Data Fig. 5**) show clear localization of these transcripts in excitatory neurons of the superficial cortical layers (*CUX2* staining). Quantification of transcript abundance (using dotdotdot^40^) confirmed the highest expression for schizophrenia individuals, a lower expression for control individuals, and the lowest expression for transcriptionally-resilient individuals for *TCF4* and *CLU*, and the opposite trend for *SHANK2* and *UNC13A*, strongly confirming the cell-type-specificity, differential expression, and directionality of our findings, and also confirming the surprising behavior of transcriptionally-resilient individuals.

### Dysregulated genes converge on synaptic plasticity, organization, and development

Observed gene expression changes suggest that schizophrenia predominantly impacts the transcriptional state of neuronal populations. To investigate whether neuronal processes are consistently dysregulated, or, alternatively, diverse neuronal functions are perturbed across cell subpopulations more specifically, we designed and performed extensive pathway enrichment analyses. We curated and organized relevant biological pathways into 14 neuronally-associated functional categories within three major themes with direct relevance to the etiology of schizophrenia: synaptic organization^41^, synaptic plasticity^2^, and neurodevelopment^42^ (**Supplementary Table 8a**). We observe a pan-neuronal overrepresentation of pathways related to post-synaptic processes within both up- and downregulated genes, consistent with genetic and proteomic evidence that schizophrenia perturbations converge on the postsynaptic density^43,44^. Neurodevelopmental pathways show higher enrichment among upregulated genes, whereas glutamate signaling and synaptic plasticity are dominantly downregulated in schizophrenia. Among subpopulations of inhibitory neurons, PV- and SST-expressing neurons show the most robust functional enrichment with up- and downregulated genes, respectively (**Fig. 2e, Supplementary Table 8b**,**c**).

In addition to these major categories, we identified the disruption of cytoskeletal processes to be specifically enriched in the Ex-SZTR cell-state. This includes differential expression of genes involved in lamellipodium organization and assembly, migration (*CAPZB, CARMIL1, SLIT2*), morphologic regulation of axons, dendrites, and dendritic spines (*CARMIL1, GOLPH3, SRGAP2, VAV2*^*45*^, *VAV3, WASF3*^*46*^), and axon guidance (*ABLIM1, NCK1*^*47*^, *PTPRO, SLIT2*). These findings in the Ex-SZTR cell-state, which is predominantly associated with markers of the superficial cortical layers, are consistent with previous studies describing aberrant pyramidal cell density and morphology predominantly within layers II and III^48–50^.

### Correlated patterns of genetic and cell-type-specific expression perturbations

Seeking insights into the association between transcriptional alterations and schizophrenia manifestation, we investigated the relationship between dysregulated genes and genetic risk factors. By assessing the cell-type-specific differential expression of genes within 145 genomic loci significantly associated with schizophrenia^14^, we discovered significant differential expression events in at least one cell type within 68 of these loci (**Fig. 3a, Supplementary Table 9)**. These novel associations identify putative causal mechanisms for more schizophrenia risk loci than previously possible using bulk tissue gene expression data, explaining the functional relevance of these loci with candidate genes, cell-type of action, and direction of effect.

**Figure 3.**
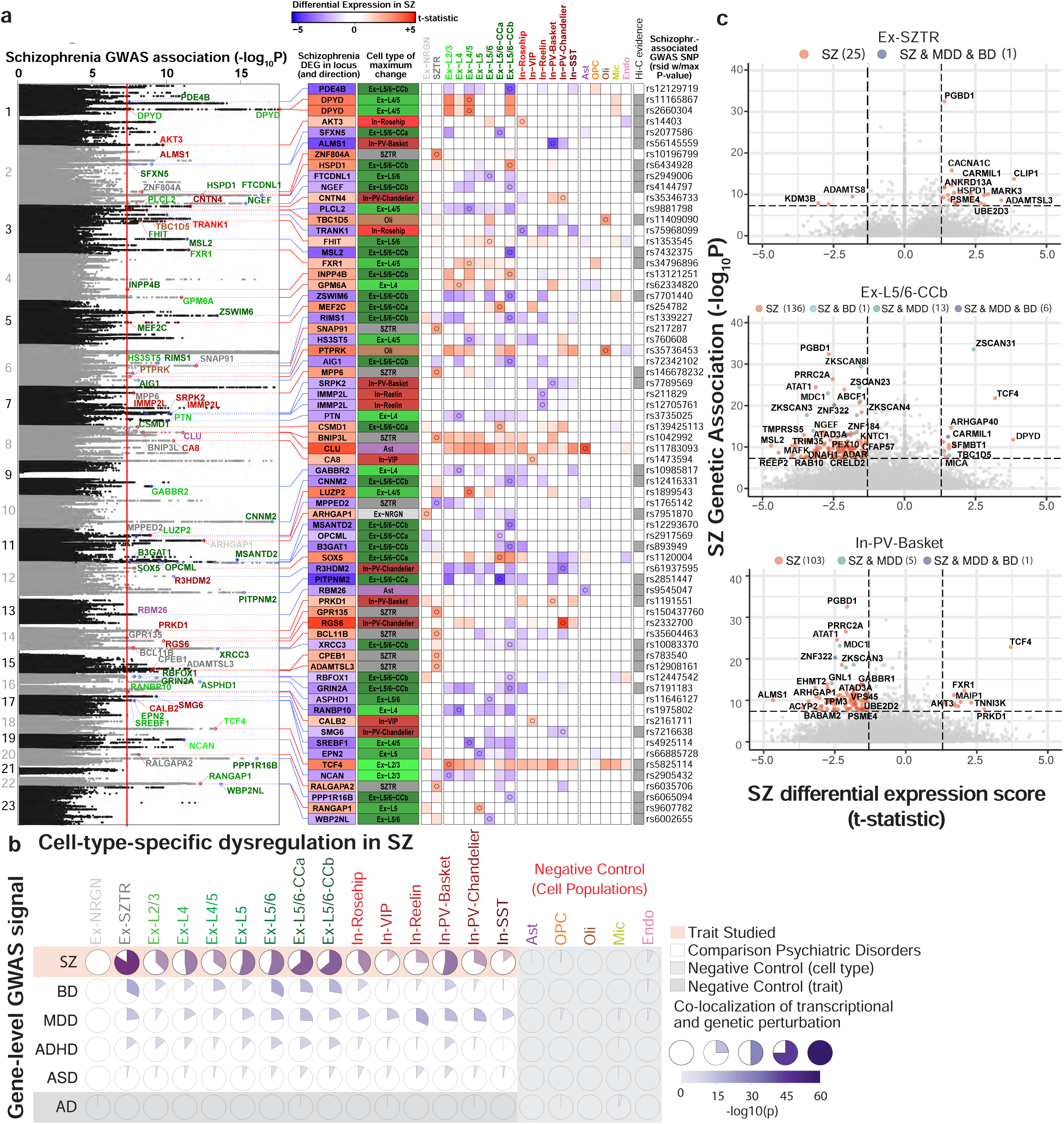
Correlation of differentially expressed genes with GWAS-associated genesets. **a**. Schizophrenia DEGs suggest mechanisms of action for 68 of 145 GWAS implicated loci. Shown are the 68 GWAS loci^14^ containing significant SZ DEGs, along with the most perturbed gene, the cell-type in which the largest DE event occurs, and a heatmap depicting all DE events for that gene within neuronal populations (red - upregulated; blue - downregulated, circles mark the maximum differential expression event for each gene). **b**. Correlation plots depicting the overlap between cell-type/state-specific schizophrenia DEGs and genes implicated by Genome-Wide Association Studies of six distinct neuropsychiatric disorders. Within each pairwise association, the ratio of purple clock-face indicates the relative amount of overlap, with the intensity of coloring indicating the significance of the association. **c**. Visualization of the relationship between the level of significance of genome-wide association with schizophrenia (y-axis) and the magnitude of differential expression in schizophrenia (x-axis) for individual genes within three neuronal populations.

The majority of schizophrenia DEGs providing explanation for these 68 genomic loci were maximally perturbed in excitatory neuronal populations, while 13 were most robustly altered in inhibitory neurons, and two genes in astrocytes and two in oligodendrocytes. Perturbation of explanatory genes within these regions was split nearly equally between up and downregulation (32 up vs. 36 down). Many explanatory genes were dysregulated in a small number or only a single cell-type, or dysregulated in opposing directions across multiple cell-types, resulting in their not being identified as schizophrenia DEGs in prior studies of bulk tissue. For example, the *CALB2* gene encodes the calcium binding protein calretinin which is expressed specifically in In-VIP and In-SST populations and contributes to long-term potentiation through regulation of neuronal excitability^51^, and *CALB2* was specifically upregulated in only the In-VIP cell-type. *ALMS1* encodes a microtubule organizing protein, and this gene was downregulated specifically in PV-expressing interneurons. Similarly, we found multiple GWAS explanatory genes that were specifically dysregulated in the Ex-SZTR cell-type (*CPEB1, GPR135, ZNF804A*), or upregulated in Ex-SZTR but downregulated across other populations (*BCL11B, RALGAPA2*).

In addition to schizophrenia^14^, we further considered four psychiatric disorders that are known to share genetic risk factors^52^: major depressive disorder (MDD)^53^, bipolar disorder (BD)^54^, autism spectrum disorder (ASD)^55^, and attention-deficit/hyperactivity disorder (ADHD)^56^. This approach allows us to distinguish schizophrenia from general psychiatric illness-related associations. As a point of contrast, we included Alzheimer’s disease (AD)^57^, a neurodegenerative disorder not expected to be genetically related to schizophrenia. For each gene/trait pair, we computed an aggregate genetic perturbation score using H-MAGMA^52^, a tool that associates GWAS risk variants to genes based on proximity to gene bodies, promoter regions, or regions linked to genes through chromatin looping. We only considered evidence of distal regulatory links occurring in the adult dorsolateral prefrontal cortex^26^. As a measure of transcriptional perturbation for schizophrenia, we used the absolute value of the t-statistics computed in the cell-type-specific differential expression analysis.

Correlation analysis between genetic and transcriptional perturbation scores for schizophrenia uncovered a strong association within neuronal subpopulations, suggesting a substantial causal effect for observed transcriptional alterations (**Fig. 3b**). We found that many of the cell-types whose transcriptional perturbations show a strong association with schizophrenia genetic risk variants are also associated with risk variants for bipolar and major depressive disorders, which is consistent with their previously-reported strong genetic relationships both at the level of genetic correlations and gene-level overlaps^52^. We found weaker relationships between schizophrenia transcriptional perturbations and ASD risk^52,58^, consistent with the known lower correlation of genetic risk between these disorders^58^. Indeed, schizophrenia transcriptional perturbations showed overlap with genetic risk loci for all neuropsychiatric disorders we assessed, while as expected we found no overlap with genetic risk for the neurodegenerative disorder AD.

Across excitatory neuronal subpopulations, the relation between transcriptional and genetic perturbation was strongest for the Ex-SZTR cell state, followed by deep-layer corticocortical projection neurons. The Ex-SZTR cell state is predominantly associated with superficial-layer excitatory neurons (layers II/III), whereas cortico-cortical excitatory neurons are enriched for layer V. The observed layer-specificity of perturbations is consistent with the enrichment of schizophrenia genetic variants proximal to genes preferentially expressed in layer II and layer V^59^. Among inhibitory neurons, we detected the strongest association between schizophrenia DEGs and genomic variants in In-PV-Basket cells, consistent with the known dysregulation of PV-expressing GABAergic neurons in schizophrenia hypothesized to contribute to the disruption of synchronous neural activity in schizophrenia^60,61^. Unlike neuronal populations, we did not observe any significant association between patterns of differential expression and genetic association in either glial or endothelial cells, suggesting non-neuronal cell types do not play a primary role in schizophrenia pathologies mediated by transcriptional dysregulation.

We next examined genes that show both genetic and transcriptional perturbations, focusing on the cell-types most enriched for schizophrenia DEGs (Ex-SZTR, deep-layer cortico-cortical excitatory neurons, and In-PV-Basket cells, **Fig. 3c, Extended Data Fig. 6**). Among these genes we found *TCF4*, a regulator of pyramidal cell excitability whose target genes are also schizophrenia-associated and cluster in neurodevelopmental pathways^62^; *CLIP1*, a linker protein involved in microtubule trafficking of cargo within axons and dendrites^63^; *CACNA1C*, an ion channel with key roles in synaptic plasticity and neurodevelopment; *ZKSCAN3*, a transcription factor implicated in autophagy; and *PGBD1*, a transposase with brain-specific expression and unknown functionality.

### Transcriptional dissection of trans-acting factors reveals common regulators

While individual DEGs may be affected by *cis-*acting genetic variants, coordinated expression changes are often driven by common upstream transcriptional regulators^64^. To prioritize such potential factors, we ranked a total of **1**,**632** transcription factors (TFs), based on the degree to which their putative target genes overlap with schizophrenia DEGs^65^ (**Supplementary Table 10**). From this data, we identified, for each cell type, a list of candidate regulators and tested whether their functional annotations converge to similar pathways as those observed for DEGs (**Fig. 2c**). Surprisingly, TFs were strongly enriched only in neurodevelopmental pathways (**Fig. 4a, Supplementary Table 11**), and not in synaptic function or signaling pathways.

**Figure 4.**
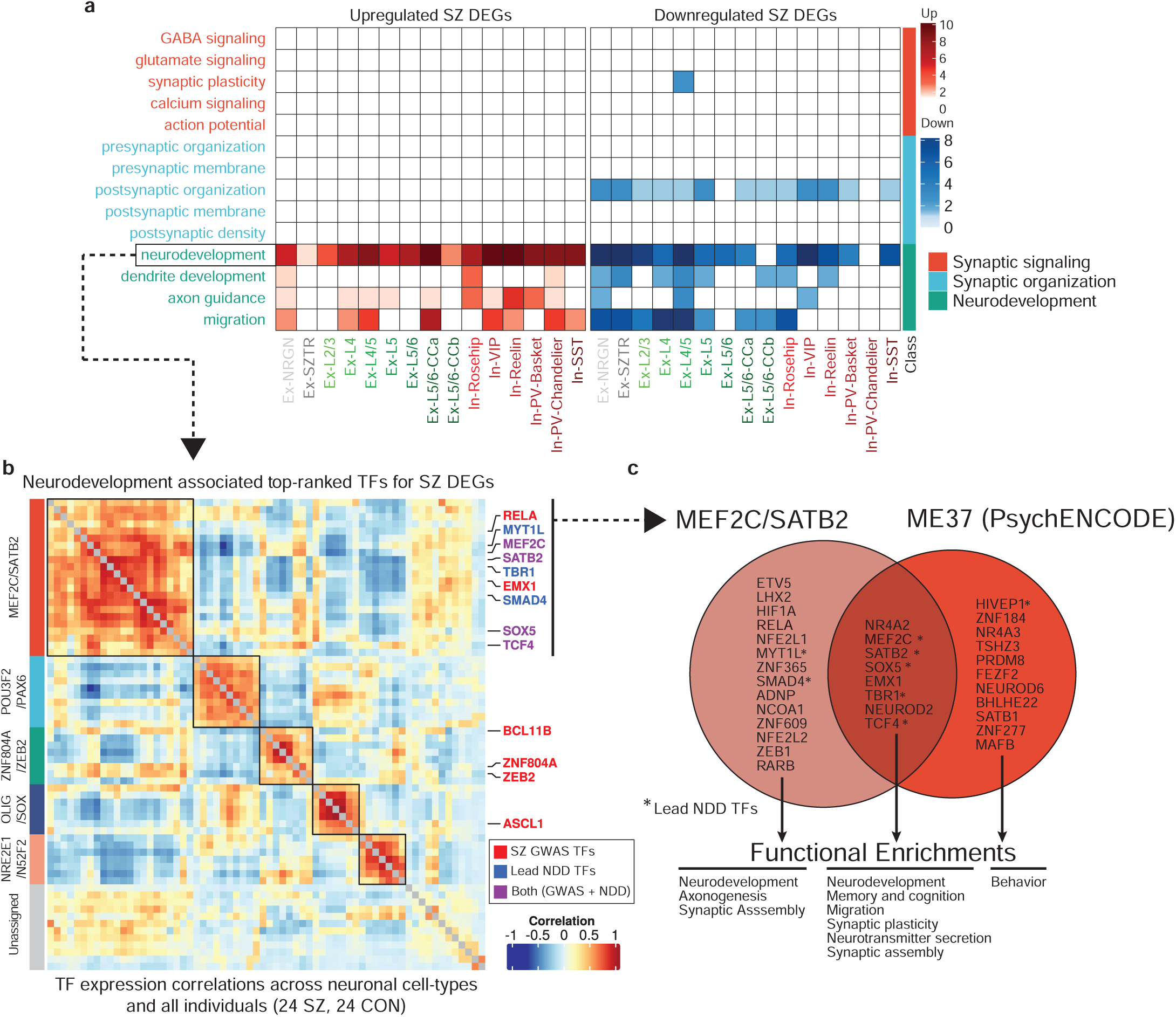
Schizophrenia-associated transcriptional regulators. **a**. Functional enrichment of the top-ranked transcriptional regulators identified by ChEA3 analysis. **b**. Co-expression heatmap of the most perturbed neurodevelopmentally-driven TFs across pseudobulk profiles from all cell-types/states and all individuals identifies five distinct TF modules, with high enrichment of GWAS-associated TFs in the MEF2C/SATB2 module (boxed top left). **c**. Overlap of TFs within the MEF2C/SATB2 module and the ME37 module identified in the PyschENCODE brain development dataset.

To assess the association of schizophrenia genetic risk loci with transcriptional regulators in addition to downstream transcriptional dysregulation directly, we focused on a subset of GWAS-associated TFs targeting DEGs. We identified a set of seven such factors (TCF4, SATB2, MEF2C, FOXG1, SOX5, ZNF536, and ZNF804A) with very strong overlap between their putative target genes and schizophrenia DEGs across all neuronal cell-types/states (**Extended Data Fig. 7)**. Interestingly, these TFs again point to neurodevelopmental functions. TCF4 is a broadly expressed helix-loop-helix transcription factor involved in nervous system development that, when disrupted, causes the severe neurodevelopmental disorder Pitt Hopkins Syndrome; SOX5 is implicated in fate determination and regulation of corticofugal projection neuron development, with loss-of-function perturbations leading to disrupted neuronal proportions and emergence timing^66^; ZNF804A is involved in neurodevelopment, neuronal migration, and protein translation. To test whether additional neurodevelopmentally-associated transcriptional regulators not implicated by schizophrenia genetic studies might be functionally relevant in the context of schizophrenia by sharing converging behavior with these lead factors, we used the set of top-ranked TFs that are also annotated with neurodevelopmental functions to identify modules of coherent TFs. We constructed a TF-TF co-expression network and applied a clustering algorithm to identify five neurodevelopmentally-related, schizophrenia-associated TF modules (**Fig. 4b, Supplementary Table 12, 13**). In addition to the lead GWAS-associated factors, we found additional factors with known neurodevelopmental roles, including TBR1, which interacts with a SOX5/SATB2 circuit thought to regulate layer specification during fetal neurogenesis^67^.

Among these modules, we found a TF group highly enriched for both schizophrenia- and neurodevelopmental delay (NDD)-associated genetic variants (**schizophrenia** p-value:1.2×10^−7^, **NDD** p-value: 3×10^−6^, Fisher’s exact test). NDD encompasses a broad category of disorders characterized by disrupted development of the nervous system leading to abnormal brain function, including intellectual disability, movement, speech, and tic disorders^68^. We observed a significant overlap between genetically-identified NDD-associated TFs (de novo mutations and CNVs)^69^ and our top-ranked schizophrenia TFs, in particular, TFs mediating upregulated genes in Ex-SZTR and Ex-NRGN, as well as downregulated genes across multiple subtypes of excitatory neurons (**Extended Data Fig. 8)**. We observed a significant over-representation of the schizophrenia-GWAS associated TFs among the top-10 ranked TFs (lead NDD TFs), where NDD-associated factors are sorted based on their overall ChEA score (*p-*value<7×10^−7^, Fisher’s exact test). The majority of the lead NDD TFs (7/10) are also involved in neurodevelopment (**Extended Data Fig. 9**). Thus, we identified a group of coregulated TFs that are associated with (1) downstream schizophrenia-associated transcriptional dysregulation in neurons of the adult cortex, (2) schizophrenia genetic risk, and (3) genetic risk for disrupted neurodevelopment.

Two of the modules identified herein are consistent with neurodevelopmentally-associated gene modules recently reported by the PsychENCODE Consortium (PEC)^70^. GWAS-enriched modules MEF2C/SATB2 and ZNF804A/ZEB2 strongly map to modules ME37 and ME32 in the PEC dataset, both of which were also reported to be enriched for schizophrenia variants. The latter module contains 19 TFs, 9 of which overlap with the MEF2C/SATB2 module (adjusted *p-*value <10^−10^, Fisher’s exact test). The overlap covers the majority of GWAS-associated TFs, including SOX5, SATB2, MEF2C, TCF4, and EMX1 (**Fig. 4c**).

### Experimentally validated TF targets confirm developmental-synaptic axis

To further investigate the link between the prioritized TFs and schizophrenia DEGs, we next tested whether active cis-regulatory elements that are targeted by these TFs show preferential association with schizophrenia DEGs. To this end, we performed Cleavage Under Targets and Tagmentation (CUT&Tag^71^) assays to map the binding of MEF2C, SATB2, SOX5, and TCF4 genome-wide in neuronal nuclei sampled from four schizophrenia and four control individuals within our larger cohort. For each transcription factor, we defined a master-set of targeted regions that are identified in at least one sample. These master-sets of regulatory elements were then projected onto the set of regulatory elements defined by the PEC developing human brain dataset^70^, which is a union of enhancers/promoters across different stages of brain development (**Extended Data Fig. 10**). Finally, we mapped these TF-centric active regulatory elements to their putative target genes, considering both proximal and distal interactions.

Given the set of target genes for each of these factors, we performed functional enrichment analysis to uncover the neuronally-associated categories that are related to each TF’s targets (**Fig. 5a**). Observed targets of these TFs show a functional enrichment profile highly similar to that of up- and downregulated schizophrenia DEGs (**Fig. 2e**), with prominent enrichment in categories related to postsynaptic and neurodevelopmental processes, supporting a functional role for these TFs in dysregulation of these events in schizophrenia.

**Figure 5.**
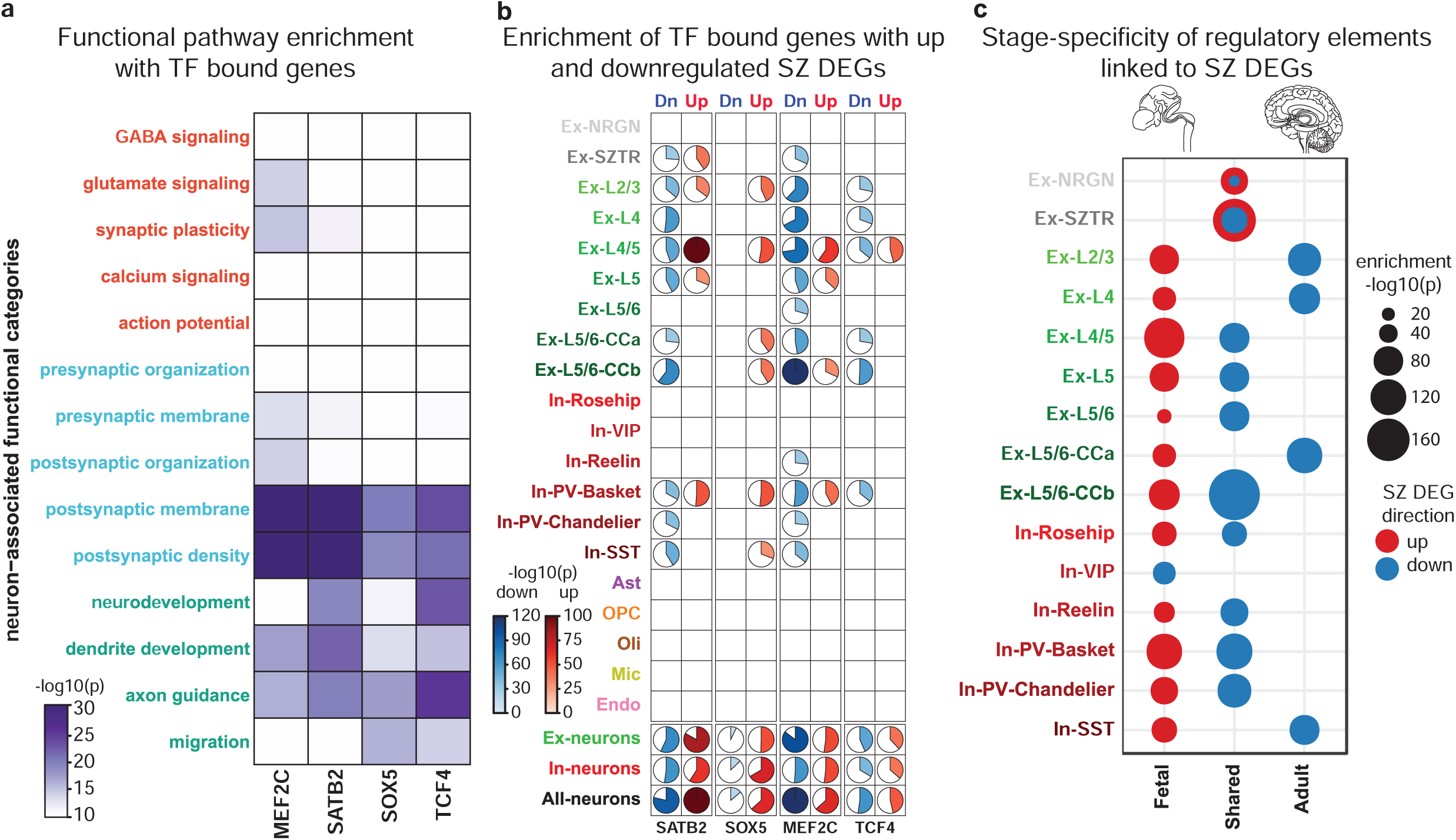
Functional characterization of TFs targeting schizophrenia DEGs. **a**. The overlap between neuronal cell-type/state-specific schizophrenia DEGs and bindings sites for MEF2C, SATB2, SOX5, and TCF4 directly assessed by CUT&Tag assays in FANS sorted neuronal nuclei from four schizophrenia and four control individuals. **b**. TF targeted genes enrich neuron-associated functional categories with a pattern highly similar to that seen for schizophrenia DEGs (Fig. 2e). **c**. Association of TFs targeting up- (red) and downregulated (blue) schizophrenia DEGs with developmental stage-specific enhancers within each neuronal cell-type/state. In all panels significance of enrichments was assessed with Fisher’s exact test.

We then assessed the overlap of target genes with up- and downregulated schizophrenia DEGs independently (**Fig. 5b**). We found a high degree of overlap between identified target genes and schizophrenia DEGs across a wide-range of excitatory neurons, with the largest overlap between TF targeted genes and Ex-L4/5 upregulated and EX-L5/6-CCb downregulated DEGs. Among inhibitory neuronal populations, PV and SST expression subtypes which originate in the medial ganglionic eminence showed markedly stronger associations than inhibitory neurons originating in the caudal ganglionic eminence (In-Reelin, In-Rosehip, and In-VIP). Finally, we did not observe a strong association between identified target genes and DEGs in non-neuronal cell-types.

Target genes for SOX5 and SATB2, which play prominent roles in early neurodevelopment, exhibit a higher degree of overlap with upregulated genes than MEF2C or TCF4. Conversely, SOX5 has an insignificant association with downregulated genes, whereas MEF2C shows the strongest association with downregulated genes, consistent with MEF2C’s role as a transcriptional repressor^72^. This suggestion of up and downregulated schizophrenia DEGs having distinct associations with neurodevelopmental stages prompted us to investigate how the regulatory landscape of schizophrenia DEGs varies across brain development. We identified cis-regulatory elements associated with different stages of brain development based on H3K27ac-enriched genomic regions profiled in the human brain by the PEC^70^, and linked enhancers with genes using evidence of physical chromatin interaction (HiC profiles) and genomic proximity. Associating fetal and adult regulatory elements independently with schizophrenia DEGs, we found that upregulated genes are predominantly linked to enhancers that are specific to the fetal stage, whereas downregulated genes are linked to enhancers that are either shared between the fetal and adult brain or are unique to the adult brain (**Fig. 5c**). Among the strongest connections, we found Ex-SZTR upregulated and Ex-L5/6-CCb downregulated genes to be associated with shared enhancers, while Ex-L4/5 upregulated genes are enriched for fetal-specific enhancers.

## Discussion

In this work, we presented the first single-cell transcriptomic case-control dissection of schizophrenia, producing an invaluable high-resolution and high-quality dataset of more than 500,000 single-cell transcriptomes. We annotated 18 neuronal and glial cell types and two excitatory-neuron cross-cutting cellular states, which we used to investigate cell-type-specific schizophrenia-dysregulated genes, pathways, and regulators. The vast majority of differentially-expressed genes were cell-type-specific and undetectable in bulk datasets, including 387 genes that were both upregulated and downregulated in distinct cell populations, highlighting the importance of our single-cell datasets.

Within neuronal subpopulations, we uncovered both wide-spread and subtype-specific transcriptional changes in both glutamatergic and GABAergic neurons, in particular PV-expressing interneurons, that are not readily detectable in bulk tissue. Our results reveal the central role in schizophrenia for genes and regulators involved in synapse formation/structure/function, with synaptic dysregulation of different neuronal subtypes implicating distinct but functionally-related genes. Unexpectedly, the transcription factors targeting these synapse-related genes enrich primarily neurodevelopmental processes, linking disruption of early neurodevelopment and adult synaptic dysfunction and providing a novel bridge between two prominent theories of schizophrenia pathogenesis. Furthermore, across multiple neuronal populations, upregulated genes implicated fetal enhancers while downregulated genes implicated adult enhancers, suggesting impairment of epigenomic differentiation of brain-specific enhancers across development as an underlying mechanistic paradigm.

Cutting across multiple subtypes of excitatory neurons within multiple layers, we uncovered a new cellular state, Ex-SZTR, overrepresented in schizophrenic individuals but surprisingly associated with transcriptional signatures of non-schizophrenia individuals across all other neuronal cell types, indicative of transcriptional resilience. This relationship between gene-regulatory changes in only one cellular sub-state and global transcriptional dysregulation in other neuronal cell types is reminiscent of the role of PV-expressing inhibitory neurons in regulating synchronous firing of assemblies of excitatory neurons, a process implicated in multiple neurodegenerative and psychiatric disorders^73,74^. Indeed, we found that PV interneurons showed the strongest correlation with non-schizophrenia (“resilience”) transcriptional signatures for the most transcriptionally-resilient individuals, suggesting a potential interplay between Ex-SZTR and PV-expressing interneurons. While the genetic and environmental factors influencing the Ex-SZTR cell state require further investigation, these tantalizing findings suggest a new potential mechanism of resilience to schizophrenia molecular pathologies that may be exploitable for new whole-brain therapeutic interventions against schizophrenia.

Our cell-type-specific DEGs showed a highly-significant enrichment in genetic loci associated with schizophrenia, with schizophrenia-differentially-expressed genes in approximately half of all schizophrenia GWAS loci^14^, including many loci previously lacking any mechanistic hypothesis. This substantial overlap suggests potential mediating roles of these genes in linking genetic causation and molecular phenotypic manifestation and helps reveal putative target genes that may be mediating the genetic effects of these loci, the cell type in which these genes may be acting, and the directionality of their effect to distinguish protective vs. risk-increasing gene expression changes. These insights can be invaluable in guiding the development of therapeutic interventions, deciding on genes to target, the cell types in which to observe their impact, and whether pharmaceutical interventions should be inhibitory or inducing, especially given the greatly-increased success of therapeutic efforts with genetic support^75^. Across GWAS loci, deep-layer excitatory neurons, cortico-cortical projection neurons, and PV-expressing basket interneurons showed the strongest enrichments, indicating central roles in mediating causal genetic effects. These enrichments between our differentially-expressed genes and GWAS loci held across multiple psychiatric disorders, consistent with the shared heritability between them^58^, but were absent from Alzheimer’s GWAS loci, providing confidence about the specificity of our analyses.

These high-resolution cell-type-specific dysregulated gene sets also enabled insights into the upstream transcription factors (TFs) most likely to influence these changes. Unlike dysregulated genes, these factors are predominantly enriched for neurodevelopmental pathways. In particular, multiple lines of evidence reveal a central role for four master regulators, namely TCF4, MEF2C, SOX5, and SATB2, which: (1) regulate schizophrenia dysregulated genes primarily in neurons; (2) are genetically associated with both schizophrenia and NDD; and (3) are key neurodevelopmental regulatory factors^70^. We experimentally validated that the target genes of these factors are schizophrenia DEGs, confirming their relevance to schizophrenia pathology. Because these TFs are major regulators of neurodevelopment with strong evidence for their action as upstream regulators of neuronal schizophrenia DEGs in adult synaptic function, we propose that their pleiotropic roles may represent a link between early neurodevelopmental disruptions and adult brain function. How genetic or early environmental perturbations to these TFs impact function to increase the risk for schizophrenia, and why it is only later in life that the phenotypic effects manifest, cannot be answered at present. However, our observations open up opportunities for future mechanistic studies tracking the developmental consequences of direct genetic perturbations to the candidate schizophrenia regulators and target genes put forward here.

While providing numerous biological insights, we acknowledge that the scope of this work is limited by current technologies. As is necessary for investigating postmortem brain tissue, we measured transcriptomes of isolated single-nuclei, and the differences between nuclear and whole-tissue RNA content must be considered in the interpretation of this work. Additionally, there are many mechanisms critical to brain function not visible to this methodology, such as mRNA splicing or trafficking of resident dendritic mRNAs. Another technical limitation of this work is the inability to spatially resolve differential expression events, and we have addressed this problem computationally and histologically where possible. These technology-specific limitations provide an opportunity for future research when newer, more advanced technologies are routinely available. Additionally, while our case-control design is a strength of the study, this focus on patient tissue does not allow experimental manipulation to investigate the causality of our validated observations, and experiments in model systems are needed. Individuals with schizophrenia have very diverse experiences of treatment and clinical outcomes. Our study design is well powered for comparison of schizophrenia vs. control, but identification of trends within subgroups of schizophrenia subjects affected by specific environmental, genetic, and treatment factors will require larger numbers of subjects.

This work utilized a rigorous study design and analysis plan to address potential confounds that are common to investigations of postmortem human brain tissue. Through balanced inclusion of schizophrenia and control subjects in each multiplexed sample preparation and sequencing library, we observed a remarkably low batch effect in our data, and almost no doublet contamination, two common problems with microfluidics-based snRNA-seq. We balanced diagnostic groups for demographic variables, including age, gender, and postmortem interval, and also controlled for these variables by including them as covariates in our analyses. We also controlled for psychiatric medications by including them as quantitative covariates in our analysis, as nearly all chronic schizophrenia patients receive longstanding pharmacologic treatment while individuals without psychiatric illness do not, and thus psychiatric diagnostic groups are not easily balanced for psychiatric medication exposures.

The data presented here offer a cell-type-specific reframing of schizophrenia transcriptional pathology by revealing specific cell populations impacted by schizophrenia genes, variants, and regulators. Identification of pleiotropic transcriptional regulators linking developmental and adult schizophrenia-associated pathologies, and the newly discovered transcriptionally-resilient cell state, provide avenues for future research to unravel the genetic and environmental underpinnings of this complex and heterogeneous disease, with promise for improving outcomes and quality of life for patients and their families.

## Materials and methods

### Assembly of the Tissue Cohort

We obtained postmortem human Brodmann Area 10 tissue from 24 schizophrenia subjects and 24 control individuals matched for age, gender, and postmortem interval from the Harvard Brain Tissue Resource Center at McLean Hospital. Institutional review board approval was obtained by the Harvard Brain Tissue Resource Center for the collection, storage, and distribution of brain tissue and de-identified clinical information for each case. Each case was assigned a consensus diagnosis by two psychiatrists based on a review of medical records and a questionnaire completed by the donor’s family. All cases were examined histologically, and cases with neuropathology diagnosed by tissue examination were excluded. Cases were obtained through family referral, and no cases were referred by a medical examiner’s office. Demographic variables for the assembled cohort are listed in Table S1. Upon arrival at the Harvard Brain Tissue Resource Center, fresh brains were dissected, and Brodmann Area 10 tissue was identified and quickly frozen with liquid nitrogen vapor prior to storage at −80°C.

### Assessment of Medication Exposures

To control for the effects of psychotropic medications on disease-associated changes in gene expression, medical records were reviewed and each subject’s history of medication exposure was compiled for inclusion in our differential gene expression analysis. Medications were clustered into categories including selective serotonin reuptake inhibitors, tricyclic antidepressants, mood stabilizers, antiepileptic drugs, benzodiazepines, and neuroleptics. Neuroleptic drugs were further classified by structural category, including benzisoxazoles, butyrophenones, dibenzodiazepines, indoles, phenothiazines, phenylpiperazines, thienobenzodiazepines, and thiothixenes. Within each medication category, the total number of distinct agents prescribed to each subject at any time was input to the linear model (**Supplementary Table 1**).

### Sample processing and single-cell RNA sequencing

A Nissle stained cryosection from each tissue block was examined microscopically to verify that sampled tissue included all six cortical layers and underlying white matter. On dry ice 50 mg of tissue was cut from the original block and stored at −80°C until further use. Tissue samples were then processed in batches of nine (three schizophrenia, three control, and three samples not analyzed in the current study) using a protocol adapted from a previous study^22^. Tissue was thawed in 0.5 ml ice-cold homogenization buffer (320 mM sucrose, 5 mM CaCl2, 3 mM Mg(CH3COO)2, 10 mM Tris HCl pH 7.8, 0.1 mM EDTA pH 8.0, 0.1% IGEPAL CA-630, 1 mM β-mercaptoethanol, and 0.4 U μl−1 recombinant RNase inhibitor (Clontech)) prior to homogenization with 12 strokes in a 2 ml Wheaton Dounce tissue grinder. Tissue homogenates were passed through a 40 µm cell strainer, mixed with an equal volume of working solution (50% OptiPrep density gradient medium (Sigma-Aldrich), 5 mM CaCl2, 3 mM Mg(CH3COO)2, 10 mM Tris HCl pH 7.8, 0.1 mM EDTA pH 8.0, and 1 mM β-mercaptoethanol), layered on top of an Optiprep density gradient (750 µl 30% Optiprep solution (30% OptiPrep density gradient medium,134 mM sucrose, 5 mM CaCl2, 3 mM Mg(CH3COO)2, 10 mM Tris HCl pH 7.8, 0.1 mM EDTA pH 8.0, 1 mM β-mercaptoethanol, 0.04% IGEPAL CA-630, and 0.17 U μl−1 recombinant RNase inhibitor) on top of 300 µl 40% optiprep solution (35% OptiPrep density gradient medium, 96 mM sucrose, 5 mM CaCl2, 3 mM Mg(CH3COO)2, 10 mM Tris HCl pH 7.8, 0.1 mM EDTA pH 8.0, 1 mM β-mercaptoethanol, 0.03% IGEPAL CA-630, and 0.12 U μl−1 recombinant RNase inhibitor)) and centrifuged at 10,000 g for 10 minutes at 4°C. Nuclei were recovered from the gradient, and resuspended in an equal volume of 1% BSA in phosphate buffered saline (PBS) prior to labeling with sample-specific cholesterol conjugated oligonucleotide hashtags (Integrated DNA Technologies). Labeled nuclei were washed with 1% BSA in PBS and pelleted by centrifugation at 500 g at 4°C for 5 minutes for three washes. After the final wash, nuclei were counted on a hemocytometer, and equal numbers from each sample were combined. The pooled nuclei were then applied to all eight channels of one 10x Genomics Chip B, targeting recovery of 20,000 nuclei per channel. 10x Genomics and hashtag libraries were prepared as per standard 10x Genomics Chromium Single Cell 3’ Reagent Kits v.3 and MULTI-Seq^27^ protocols.

Libraries were sequenced in batches of two on an Illumina NextSeq500 instrument (two snRNA-Seq libraries and two hashtag libraries per flowcell - average 3.6E7 reads per hashtag library), and an additional round of sequencing was performed for all snRNA-Seq libraries on an Illumina NovaSeq instrument (eight or 16 libraries per NovaSeq S2 flowcell) to achieve an average sequencing depth of 35,142 reads per cell. Gene count matrices were generated by aligning reads (including intronic reads) to the hg38 genome using 10x Genomics Cell Ranger software v3.0.1 (**Supplementary Table 2**).

### Cross-individual doublet-detection using sample hashtags

The deMULTIplex R package^27^ was used to process hashtag FASTQ files, extracting 10x barcode, sample hashtag, and UMI information for each read. Duplicated UMI and mismatched hashtag reads were excluded and retained reads were converted to a 10x barcode by sample hashtag count matrix. This count matrix was processed with the Seurat R package^76^ using the HTODemux function to cluster cells based on sample hashtag counts and determine a count threshold for each hashtag based on a negative binomial distribution applied to the cluster with the lowest expression for that hashtag. This threshold identified each cell as positive or negative for each sample hashtag, and cells identified as positive for more than one hashtag were assigned as inter-sample doublets and removed from the study (**Supplementary Table 2**).

### Characterization, visualization, and annotation of cell subpopulations using ACTIONet

Filtered data after doublet-detection was used as input to the archetypal analysis for cell-type identification (ACTION) algorithm ^77^ to identify a set of transcriptional archetypes, each representing a potential underlying cell type/state. Using ACTION-decompositions with varying numbers of archetypes, we employed our recently developed ACTION-based network (ACTIONet) framework^28^ to create a multi-resolution nearest neighbor graph. A modified version of the stochastic gradient descent-based layout method was used in the uniform manifold approximation and projection (UMAP) algorithm (Becht et al., 2018), to visualize the ACTIONet graph. ACTIONet framework identifies dominant transcriptional patterns (or archetypes) and associates cells to these archetypes with different degrees of confidence. To discretize cell associations, we assigned each cell to its most closely associated archetype. We then used a curated set of reproducible cell-type-specific markers in the human prefrontal cortex^28^ to annotate cells assigned to each archetype.

To filter cells, we performed two independent iterations of the ACTIONet framework, the first one to identify additional low-quality cells and missed doublets, and the second one to identify and annotate cell types/states. In the first stage, we removed two archetypes that were assigned to multiple unrelated marker sets, as well as cells that are not well-connected to other cells in the ACTIONet. To identify the overall connectivity of each cell within the ACTIONet graph that is assigned to a given archetype, we computed its “coreness” ^78^ within the subgraph induced by the cells associated with the same archetype. In the second round, we annotated and grouped archetypes into major cell types in the human prefrontal cortex, including subtypes of excitatory and inhibitory neurons, as well as non-neuronal cell types, as well as two novel schizophrenia-associated cell states (**Fig. 1b**,**d**). We used the coreness of cells in the ACTIONet as their transparency to de-emphasize low-quality cells. To verify our annotations, we projected individual marker genes for different subtypes on the ACTIONet (**Fig. 1c**). We did not observe any batch effect and cells from all batches and both phenotypes were well-mixed in the ACTIONet plot (**Extended Data Fig. 1b, c**), and none of the archetypes is influenced by a minority of individuals, and the majority of individuals contribute to all archetypes (**Fig. 1f**).

### Differential gene expression analysis using a modified pseudo-bulk approach

Following recent studies showing the success of pseudo-bulk methods for multi-sample multi-group differential analysis of single-cell datasets with complex experimental designs ^79^, we developed a new method based on the analysis of pseudo-bulk profiles to identify perturbed genes in schizophrenia. We aggregated the expression of genes within each archetype/individual combination and used the linear-modeling approach in the Limma package^80^ to include our experimental design in the differential analysis. However, unlike the case of bulk analysis, in which the underlying variance of each gene in each sample is unknown, in single-cell pseudo-bulk analysis we can compute both the mean and the variance of genes directly from single-cells, which provides a more accurate approach than either Limma-trend or Limma-voom^81^. We used the inverse of the gene variances as weights in our linear model and used an outlier detection approach to mask out genes that were deemed not reliable, on a per-sample basis. For each pseudo-bulk profile, we required that it has to contain at least 50 cells to be included in our model. Finally, to account for individual-specific differences, we incorporated age, gender, PMI, batch, and medication history as covariates in our model. We filtered results based on a raw p-value cut-off of 0.05 and LFR of 0.1 to declare differential genes.

### Computing transcriptional pathology scores

To assess the degree to which different cell types/states in each individual are affected by schizophrenia-associated transcriptional perturbations, we used our pseudo-bulk profiles. For each archetype, we computed a representative expression vector by averaging all pseudo-bulk samples. As a measure of schizophrenia-associated transcriptional effect in each individual/cell type, we used partial Pearson correlation between individual pseudo-bulk profiles with differential perturbation scores, after controlling for the baseline cell type/state-specific representative vector. We then scored and ranked each individual scored according to their average correlation across all archetypes.

### Identification of transcriptional regulators using ChEA3

We used ChIP-X Enrichment Analysis 3 (ChEA3)^65^ to prioritize transcription factors (TFs) that are likely to mediate the observed transcriptional dysregulation in schizophrenia. In summary, ChEA3 integrates multiple libraries of putative TF-target lists, gathered from different sources, including TF-gene co-expression from RNA-seq studies, TF-target associations from ChIP-seq experiments, and TF-gene co-occurrence computed from crowd-submitted gene lists, to compute a composite ranked list of TFs. We used the *TopRank* strategy to combine rankings from individual libraries, in which the best scaled-rank of each TF across all libraries is used to aggregate TF scores. We report the log-transformed scaled-rank as the relevance measure of each TF by ChEA3 analysis.

### Gene-centric analysis of common variants using H-MAGMA

Hi-C-coupled MAGMA (H-MAGMA) is a recent extension of the traditional multimarker analysis of genomic annotation (MAGMA), a method developed to prioritize genes by aggregating single nucleotide polymorphism associations to nearest genes. In H-MAGMA, the linking of SNPs to genes is extended to include long-range interactions brought together through the chromatin looping. In our analysis, we used the preprocessed dataset of SNPs-gene links based on the Hi-C data in the adult human brain.

### Fluorescence *in situ* hybridization (FISH)

From our larger cohort of postmortem human BA10 tissue, six cases were selected for FISH experiments including CON10, CON14, SZ5, SZ12, SZ20, and SZ23. Frozen tissue blocks were embedded in optimal cutting temperature medium (OCT), and 10 μm sections were cut at −20 °C with a Microm HM560 cryostat, mounted on Superfrost Plus slides (one control, one schizophrenia, and one transcriptionally-resilient case mounted on each slide to ensure balanced processing) and stored −80°C until FISH was conducted.

Advanced Cell Diagnostics (ACD) designed the *in situ* hybridization probes (human *SHANK2, UNC13A, TCF4*, and *CLU*) as well as the positive and negative control probes (**Supplementary Table 14**). The RNAscope Multiplex Fluorescent Reagent Kit v2 (ACD) was used for the assay following the manufacturer’s instructions with some modifications. Nuclei were counterstained with DAPI using TSA buffer (ACD) and TSA Plus fluorophores (PerkinElmer). Briefly, frozen sections were fixed for 1 h at 4 °C using freshly prepared ice-cold 10% neutral buffered formalin, and rinsed with phosphate buffered saline (PBS) and dehydrated in 50%, 70%, and two changes of 100% ethanol (5 min each) at room temperature (RT). Sections were air-dried and a hydrophobic barrier drawn around each section with an Immedge pen (Vector Laboratories). When completely dry, sections were treated with hydrogen peroxide for 10 min at RT, washed twice with PBS, incubated with protease IV for 15 min at RT, and washed again twice with PBS.

After diluting the probes for RNA detection 1:50, sections were hybridized with the probes (40°C, 2 h; HybEZ Hybridization System (ACD)), washed twice, and stored overnight at RT in 5x SSC buffer. The following day, slides were washed twice with the wash buffer, followed by three amplification steps (AMP 1 (30 min), AMP 2 (30 min), and AMP 3 (15 min); 40°C). Each amplification step was followed by two washes of 2 min each with the wash buffer. Sections were then incubated sequentially with the HRP reagent corresponding to each channel (e.g. HRP-C1; 40°C, 15 min) followed by the respective TSA Plus fluorophore assigned to the probe channel (Opal dyes, 520, 620 and 690, dilution 1:1500; 40°C, 30 min) and HRP blocker (40°C, 15 min), and each incubation was followed by two wash steps. Lipofuscin autofluorescence was visible in both green and red channels. Since the far-red channel showed less autofluorescence, highly expressed *UNC13A*-C4 and *CLU*-C2 probes were assigned to the green fluorescein channel and the *CUX2* layer-specific marker was assigned to the red cyanine 5 channel. DAPI (30 s) was used to visualize cell nuclei. Sections were mounted using ProLong Gold mounting medium (Thermo Fisher Scientific) and stored at 4 °C. Two independent experiments were performed, with three biological replicates each, and positive and negative control probes to test for RNA quality and background signal, respectively.

### Leica Laser Scanning Confocal Microscopy

Image acquisition, processing, and quantitation were performed blind to diagnosis. All microscopy was performed at the Microscopy Core at McLean Hospital on a Leica TCS-SP8 confocal microscope. Exposure times were set separately for each of the four channels (red for Cy5, blue for DAPI, green for FITC, and orange for Texas Red) and kept similar among the cases to enable comparison. Images at 40x and 63x magnifications were visualized as maximum intensity projections of Z-stacks at 1.5 μm intervals. Two representative fields of view were selected within cortical layers II and III as identified by positive *CUX2* staining, and Z-stacks were taken throughout the depth of single cells, with ≥40 single cells per case and six cases per target gene. Adjustment of brightness, contrast, and sharpness was done using Adobe Photoshop. Quantification of transcripts was performed in an automated unbiased manner using dotdotdot^40^. For all six cases, the dotdotdot MATLAB script (https://github.com/LieberInstitute/dotdotdot) was used to process one 40x and one 63x maximum intensity projection image, counting the number of puncta in each channel overlapping with each region of interest (nucleus) identified by DAPI staining. Data presented in figure 2e represent counts of *TCF4, CLU, SHANK2*, and *UNC13A* transcripts within individual nuclei containing at least one detected *CUX2* transcript, and excluding counts of zero.

### CUT&Tag mapping of transcription factor binding in the neuronal genome

Nuclei were isolated from the postmortem human prefrontal cortex as described above and incubated with 1:1000 diluted anti-NeuN antibody (EMD Millipore MAB377X) with 0.5% BSA in PBS at 4°C with end-over-end rotation for 45 minutes. After staining nuclei were counterstained with propidium iodide and sorted on a BD FACSAria III Cell Sorter at Harvard University’s Bauer Core Facility. 100,000 neuronal nuclei were used as input for each Cleavage Under Targets and Tagmentation (CUT&Tag) assay using rabbit primary antibodies targeting MEF2C (Abcam ab211439), SATB2 (Abcam ab92446), SOX5 (Abcam ab94396), TCF4 (ProteinTech 22337-1-AP), H3K27Ac (EMD Millipore MABE647), and mouse-anti-rabbit secondary antibody (Sigma R2655) with the Vazyme pG-Tn5 CUT&Tag kit (Cellagen Technology, San Diego) according to the manufacturer’s protocol. CUT&Tag libraries were sequenced on one NextSeq500 flow cell at the MIT BioMicroCenter. Reads were aligned to the HG38 genome, processed to bedgraph format, and analyzed with the Sparse Enrichment for CUT&Run^82^ tool with “stringent” parameters retaining the top 1% of peaks.

## Supporting information

TableS2_SequencingMetrics

TableS3_Combined_marker_tables

TableS4_CCab_scores

TableS5_Combined_marker_tables_enrichments

TableS6a_DE_up_genes

TableS6b_DE_down_genes

TableS7c_TPS_scores

TableS8a_NFP

TableS8b_DE_up_gprofiler

TableS8c_DE_down_gprofiler

TableS9_GWAS_SNP_table

TableS10_ChEA_scores

TableS11a_ChEA_TFs_enrichment_NFP

TableS11b_ChEA_TFs_up_gprofiler

TableS11c_ChEA_TFs_down_gprofiler

TableS13_Regulons

TableS14_RNAScopeProbes

TableS7ac_Pseudobulk_mean_Ex-L2-3

TableS7aa_Pseudobulk_mean_Ex-NRGN

TableS7ab_Pseudobulk_mean_Ex-SZTR

TableS7ad_Pseudobulk_mean_Ex-L4

TableS7ae_Pseudobulk_mean_Ex-L4-5

TableS7af_Pseudobulk_mean_Ex-L5

TableS7ag_Pseudobulk_mean_Ex-L5-6

TableS7ah_Pseudobulk_mean_Ex-L5-6CCa

TableS7ai_Pseudobulk_mean_Ex-L5-6CCb

TableS7aj_Pseudobulk_mean_In-Rosehip

TableS7ak_Pseudobulk_mean_In-VIP

TableS7al_Pseudobulk_mean_In-Reelin

TableS7am_Pseudobulk_mean_In-PV-Basket

TableS7an_Pseudobulk_mean_In-PV-Chandelier

TableS7ao_Pseudobulk_mean_In-SST

TableS7ap_Pseudobulk_mean_AST

TableS7aq_Pseudobulk_mean_OPC

TableS7as_Pseudobulk_mean_Mic

TableS7at_Pseudobulk_mean_Endo

TableS7ar_Pseudobulk_mean_Oli

TableS7ba_Pseudobulk_variance_Ex-NRGN

TableS7bb_Pseudobulk_variance_Ex-SZTR

TableS7bc_Pseudobulk_variance_Ex-L2-3

TableS7bd_Pseudobulk_variance_Ex-L4

TableS7be_Pseudobulk_variance_Ex-L4-5

TableS7bf_Pseudobulk_variance_Ex-L5

TableS7bg_Pseudobulk_variance_Ex-L5-6

TableS7bh_Pseudobulk_variance_Ex-L5-6CCa

TableS7bi_Pseudobulk_variance_Ex-L5-6CCb

TableS7bj_Pseudobulk_variance_In-Rosehip

TableS7bk_Pseudobulk_variance_In-VIP

TableS7bl_Pseudobulk_variance_In-Reelin

TableS7bm_Pseudobulk_variance_In-PV-Basket

TableS7bn_Pseudobulk_variance_In-PV-Chandelier

TableS7bo_Pseudobulk_variance_In-SST

TableS7bp_Pseudobulk_variance_AST

TableS7bq_Pseudobulk_variance_OPC

TableS7br_Pseudobulk_variance_Oli

TableS7bs_Pseudobulk_variance_Mic

TableS7bt_Pseudobulk_variance_Endo

TableS12_FilteredNorm_JointPseudobulk_part01

TableS12_FilteredNorm_JointPseudobulk_part02

TableS12_FilteredNorm_JointPseudobulk_part03

TableS12_FilteredNorm_JointPseudobulk_part04

TableS12_FilteredNorm_JointPseudobulk_part05

TableS12_FilteredNorm_JointPseudobulk_part06

TableS12_FilteredNorm_JointPseudobulk_part07

TableS12_FilteredNorm_JointPseudobulk_part08

TableS12_FilteredNorm_JointPseudobulk_part09

TableS12_FilteredNorm_JointPseudobulk_part10

TableS12_FilteredNorm_JointPseudobulk_part11

TableS12_FilteredNorm_JointPseudobulk_part12

TableS12_FilteredNorm_JointPseudobulk_part13

TableS12_FilteredNorm_JointPseudobulk_part14

TableS1_SubjectDemographics&Medication

## Data Availability

MULTI-seq data are available at Synapse (https://www.synapse.org/#!Synapse:syn22963646). The data are available under controlled use conditions set by human data privacy regulations, and access requires a data use agreement to ensure the anonymity of the donors of postmortem human brain tissue. A data use agreement can be pursued with SAGE, who maintains Synapse and can be downloaded from their website.

## Acknowledgements

We thank the individuals and families whose donation of human brain tissue made this work possible. This work was supported by the Wilf Family Foundations and by NIH grant K08MH109759 (W.B.R.) and by NIH grants 1U01MH119509, R01MH109978, and R01AG062335 (M.K.).

## Author Contributions

This study was designed by W.B.R., and directed and coordinated by W.B.R. and M.K. W.B.R., S.S., and D.R.T. performed the snRNA-seq experiment, S.S. performed the RNAScope experiment and W.B.R. and S.S. performed the CUT&Tag experiment. W.B.R. performed data processing and S.M. and J.D.-V. performed the computational analysis. D.R.T. and M.H. reviewed medical records under supervision of W.B.R. W.B.R., S.M., J.D.-V., and M.K. wrote the manuscript.

## Data Availability

The MULTI-seq data are available at Synapse (https://www.synapse.org/#!Synapse:syn22963646). The data are available under controlled use conditions set by human data privacy regulations, and access requires a data use agreement to ensure the anonymity of the donors of postmortem human brain tissue. A data use agreement can be pursued with SAGE, who maintains Synapse and can be downloaded from their website.

## Supplementary Tables

1. Subject demographics and medications (TableS1_SubjectDemographics&Medication.xlsx)
2. Sequencing statistics (TableS2_SequencingMetrics.xlsx)
3. Cell-type/state-specific marker genes (TableS3_Combined_marker_tables.xlsx)
4. Marker gene comparison between Ex-L5/6-CCa and Ex-L5/6-CCb (TableS4_CCab_scores.xlsx)
5. Pathways enriched in top-ranked marker genes from each cell-type/state (TableS5_Combined_marker_tables_enrichments.xlsx)
6. Up-(TableS6a_up_regulated_genes.xlsx) and down- (TableS6b_down_regulated_genes.xlsx) regulated genes in schizophrenia
7. Pseudo-bulk mean (TableS7a_Pseudobulk_mean.xlsx) and variance (TableS7b_Pseudobulk_variance.xlsx) profile per cell type/state, and computed Transcriptional Pathology Scores for each subject (TableS7c_TPS_scores.xlsx)
8. Curated set of neuro-associated functional pathways (NFP) and member genes (TableS8a_NFP.xlsx), and functional pathway enrichment of up- (TableS8b_DE_up_gprofiler.xlsx) and down- (TableS8c_DE_down_gprofiler.xlsx) regulated genes
9. Schizophrenia GWAS loci linked to DEGs (TableS9_GWAS_SNP_table.xlsx)
10. ChEA scores for association of 1632 TFs with up- and downregulated genes (TableS10_ChEA_scores.xlsx)
11. NFP enrichment of TFs associated with dysregulated genes (TableS11a_ChEA_TFs_enrichment_NFP.xlsx), and functional pathway enrichment of TFs associated with the set of up- (TableS11b_ChEA_TFs_up_gprofiler.xlsx) and down- (TableS11c_ChEA_TFs_down_gprofiler.xlsx) regulated genes
12. Filtered and normalized profile of the merged pseudo-bulk mean profiles (TableS12_FilteredNorm_JointPseudobulk.xlsx), used for TF expression correlation analysis
13. TF membership of individual regulons (TableS13_Regulons.xlsx)
14. RNAscope targets, probes, channels, and fluorophores (TableS14_RNAScopeProbes.xlsx)

**Extended Data Figure 1a.**
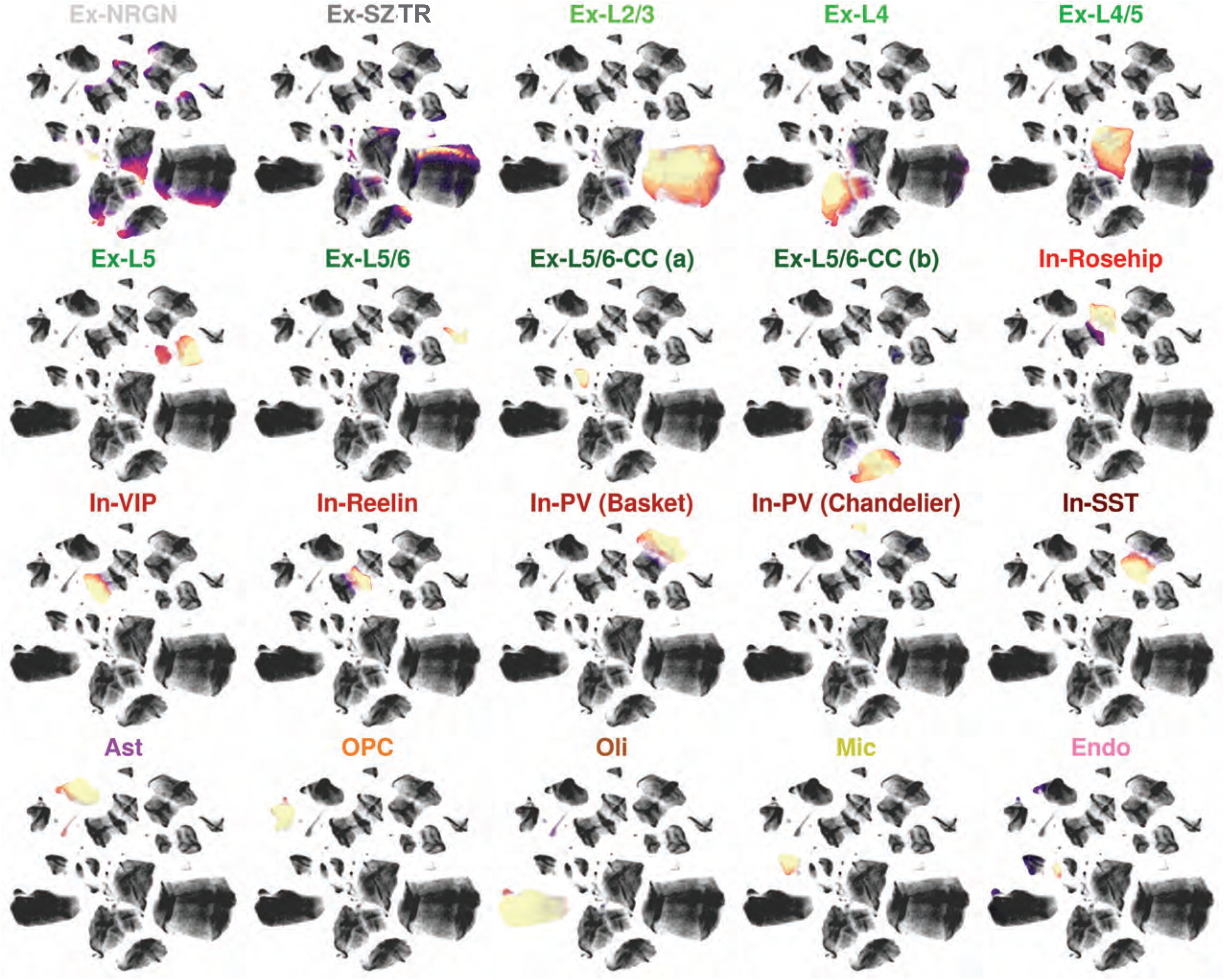
ACTIONet cell-cell similarity network depicting the footprint of all 20 identified transcriptional archetypes. **ACTIONet cell-cell similarity network**. Network-based two-dimensional visualization of all cells considered in the analysis (n=560,020) indicating association with identified transcriptional archetypes.

**Extended Data Figure 1b,c.**
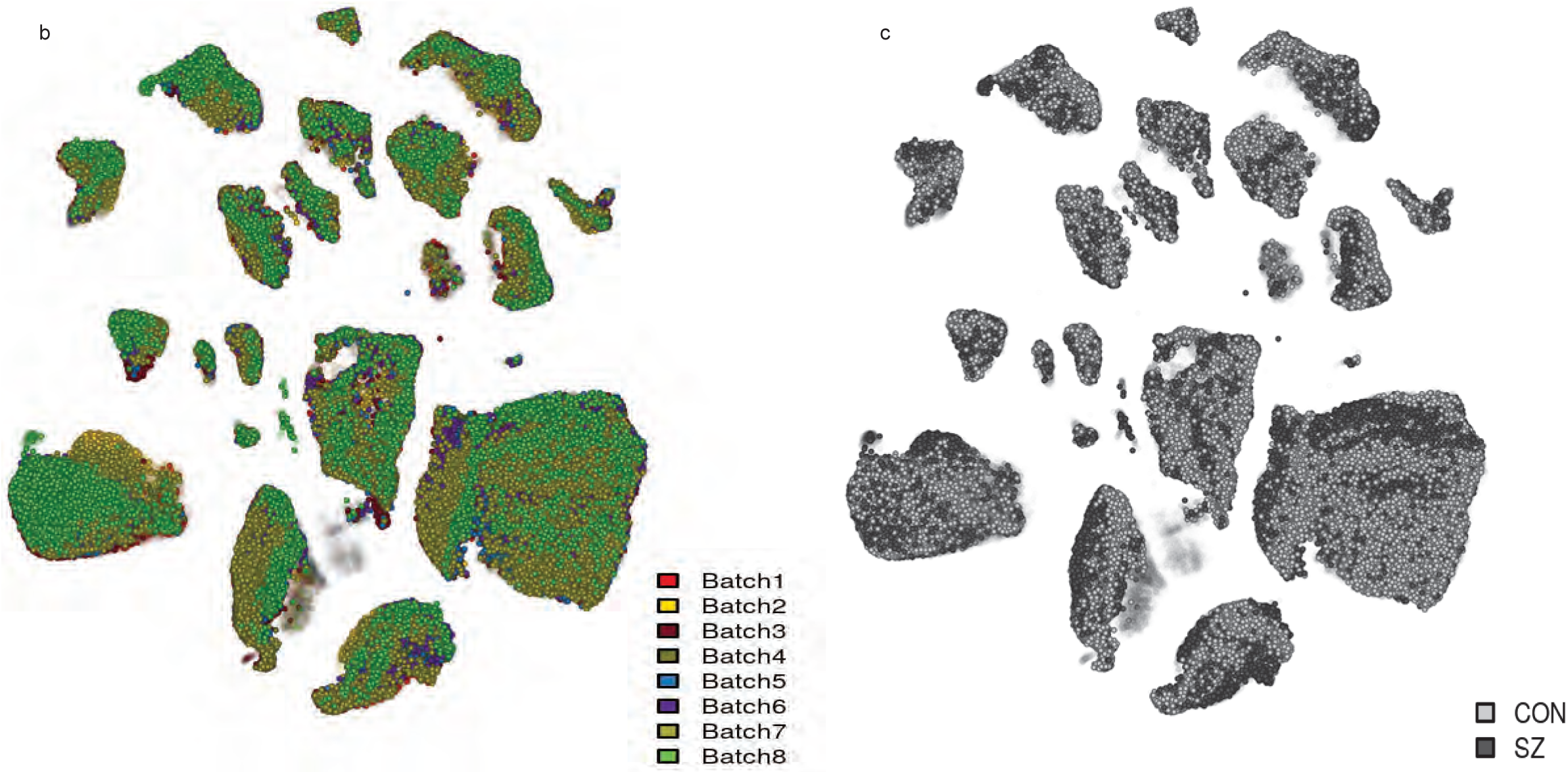
ACTIONet cell-cell similarity network colored by 10x batch (left) or phenotype (right) ACTIONet cell-cell similarity network. Network-based two-dimensional visualization of all cells considered in the analysis (n=560,020) indicating association with 10x batch (b), or phenotype (c) (SZ:schizophrenia, n=266,431; CON: control, n=293,589).

**Extended Data Figure 2a.**
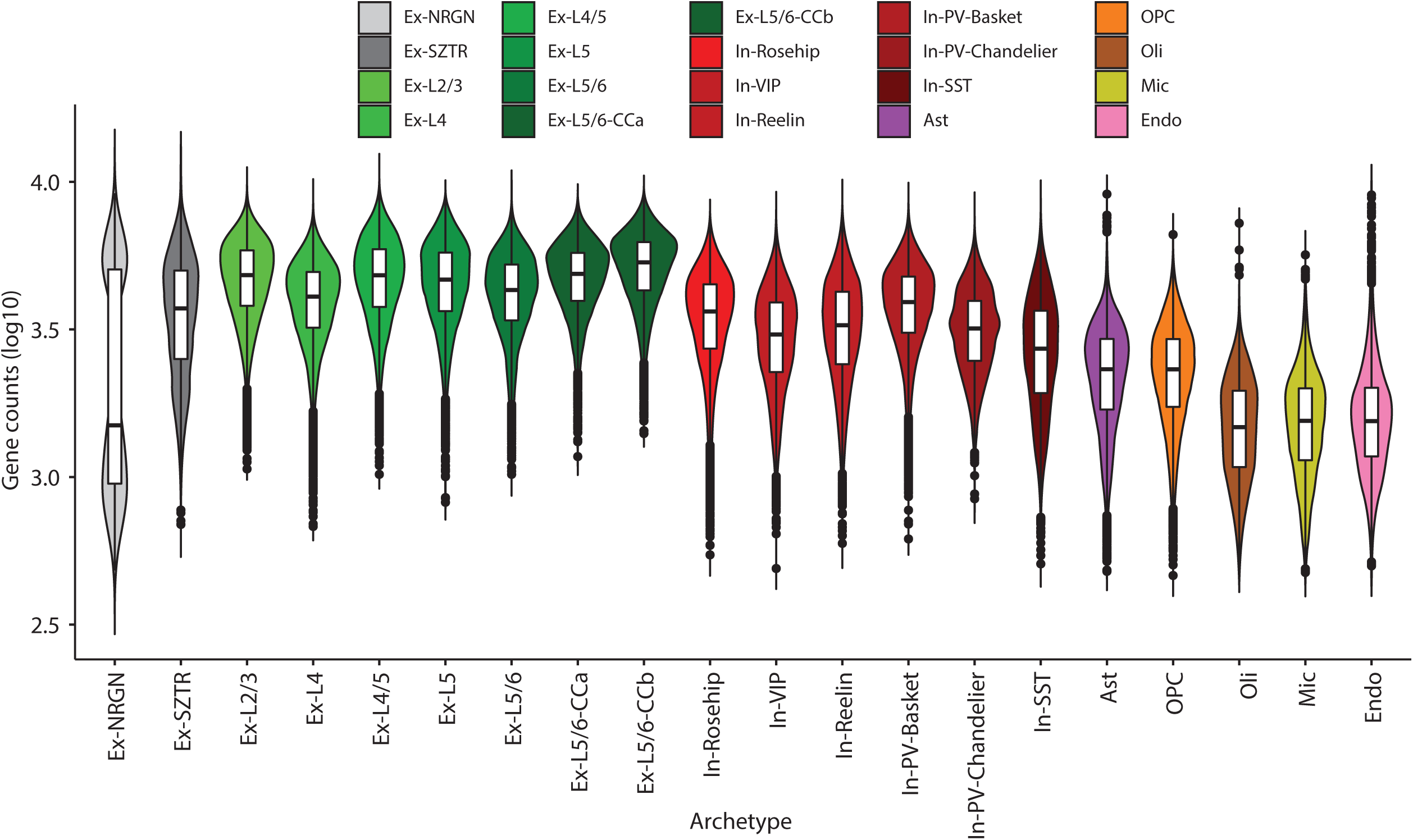
Gene capture across cell-types. **Cell type gene and cell statistics**. Gene count distribution across cells of each type. Each point represents in log scale the number of genes detected to have a read count x > 0 in a given cell. Box plots are centred around the median, with the interquartile range (IQR) defining the box. The upper whisker extends to the largest value no further than 1.5 × IQR from the end of the box. The lower whisker extends to the smallest value at most 1.5 × IQR from the end of the box.

**Extended Data Figure 2b.**
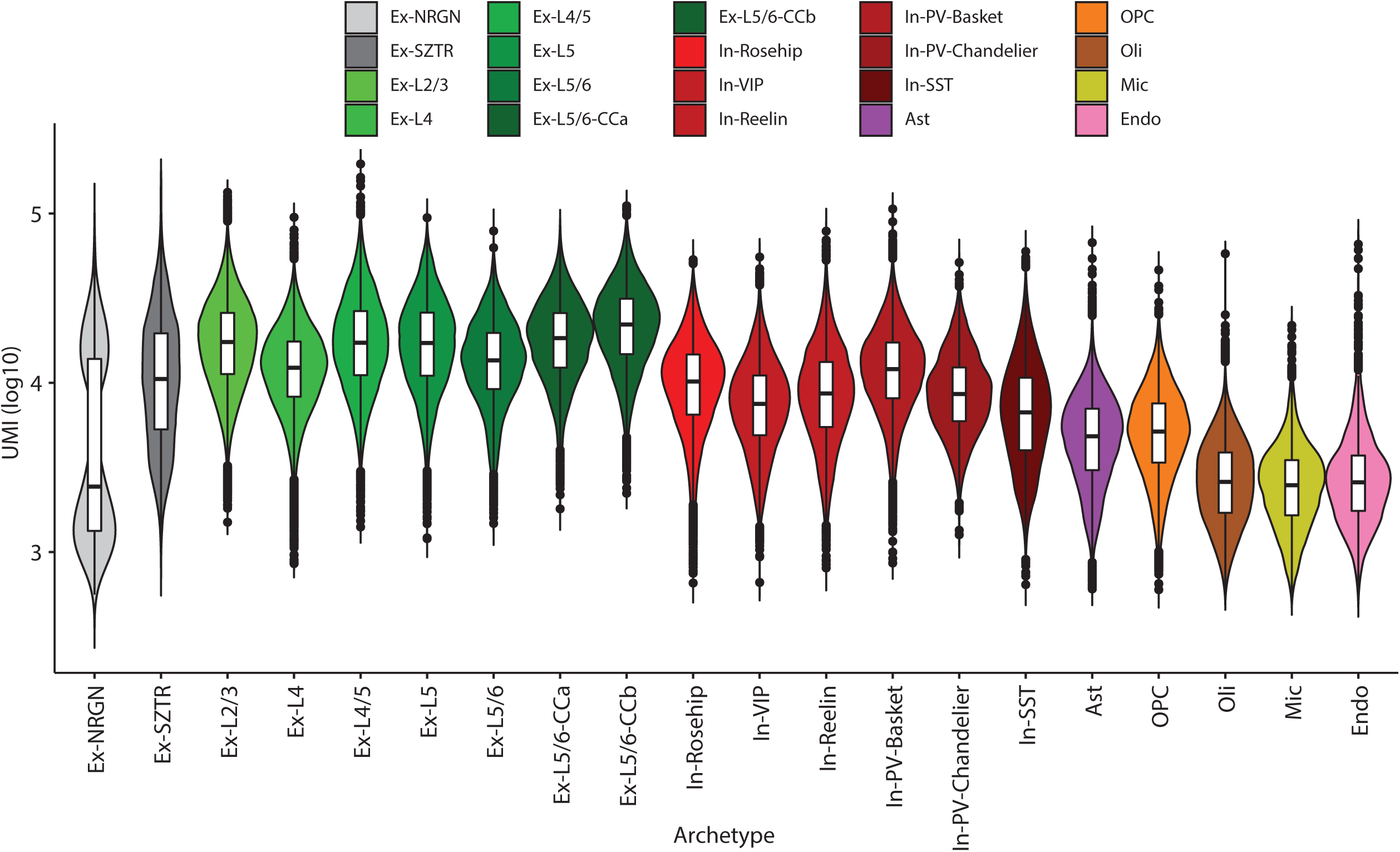
UMI capture across cell-types. **Cell type gene and cell statistics**. Total UMI count distribution across cells of each type. Each point represents the total UMI count across all genes in a given cell. Box plots are centred around the median, with the interquartile range (IQR) defining the box. The upper whisker extends to the largest value no further than 1.5 × IQR from the end of the box. The lower whisker extends to the smallest value at most 1.5 × IQR from the end of the box.

**Extended Data Figure 2c.**
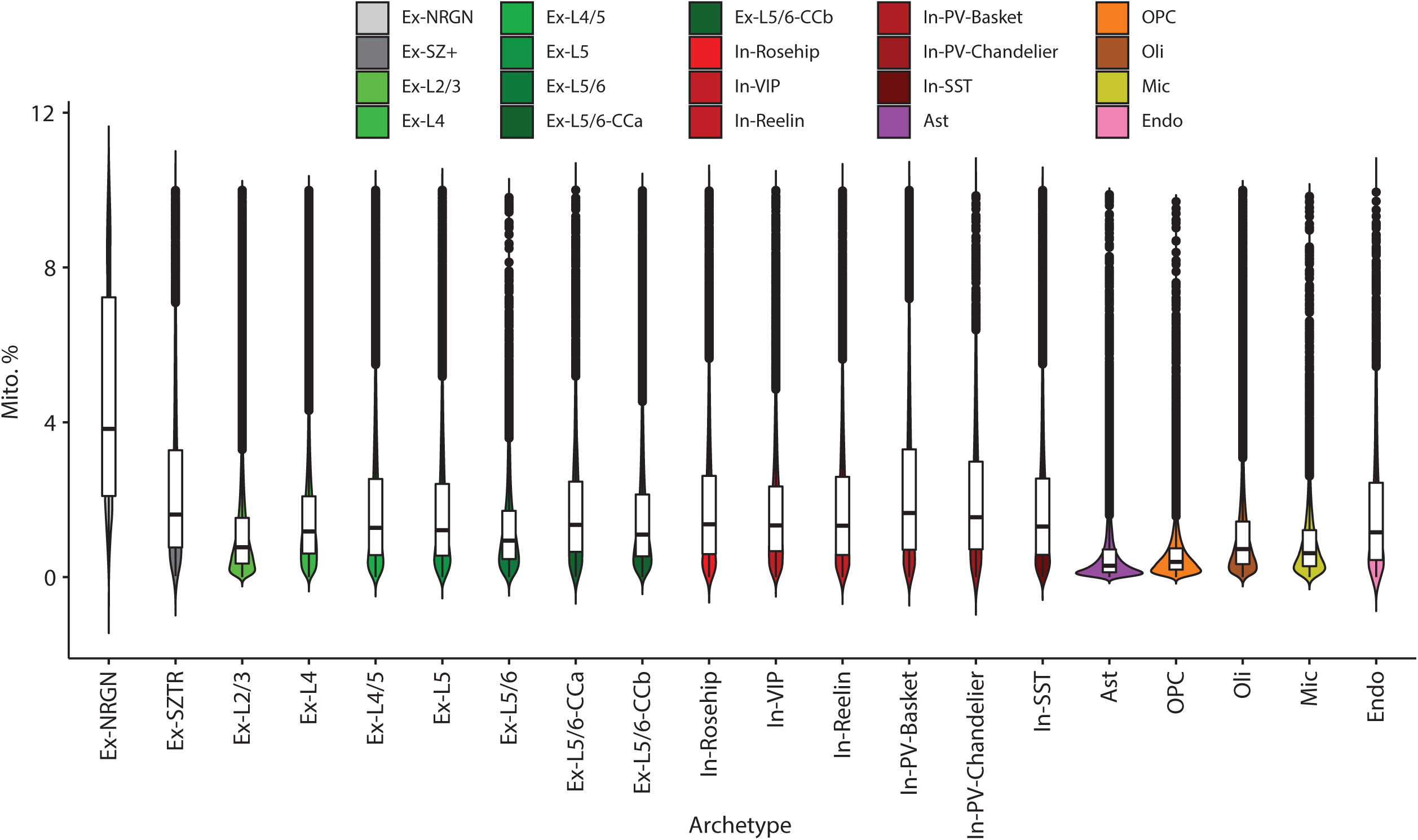
Mitochondrial gene capture across cell-types. **Cell type gene and cell statistics**. Distribution of UMI percentages that map to mitochondrially encoded genes across cells of each type. Distributions are shown for the SCZ dataset reported herein. Box plots are centred around the median, with the interquartile range (IQR) defining the box. The upper whisker extends to the largest value no further than 1.5 × IQR from the end of the box. The lower whisker extends to the smallest value at most 1.5 × IQR from the end of the box.

**Extended Data Figure 2d.**
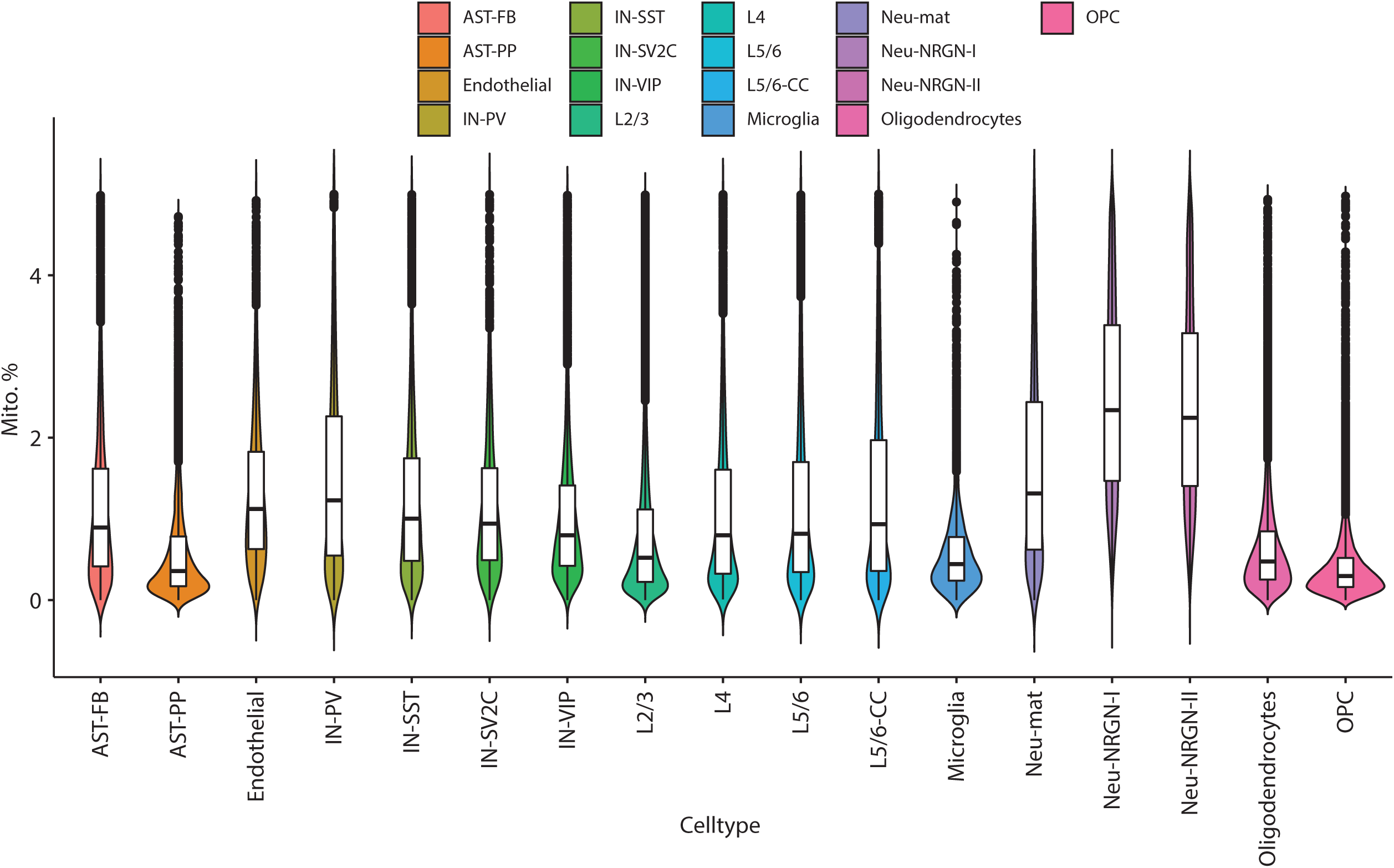
Mitochondrial gene capture across cell-types in the Velmeshev 2019 dataset. **Cell type gene and cell statistics**. Distribution of UMI percentages that map to mitochondrially encoded genes across cells of each type. Distributions are shown for the dataset reported in Velmeshev et al. 2019. Box plots are centred around the median, with the interquartile range (IQR) defining the box. The upper whisker extends to the largest value no further than 1.5 × IQR from the end of the box. The lower whisker extends to the smallest value at most 1.5 × IQR from the end of the box.

**Extended Data Figure 3.**
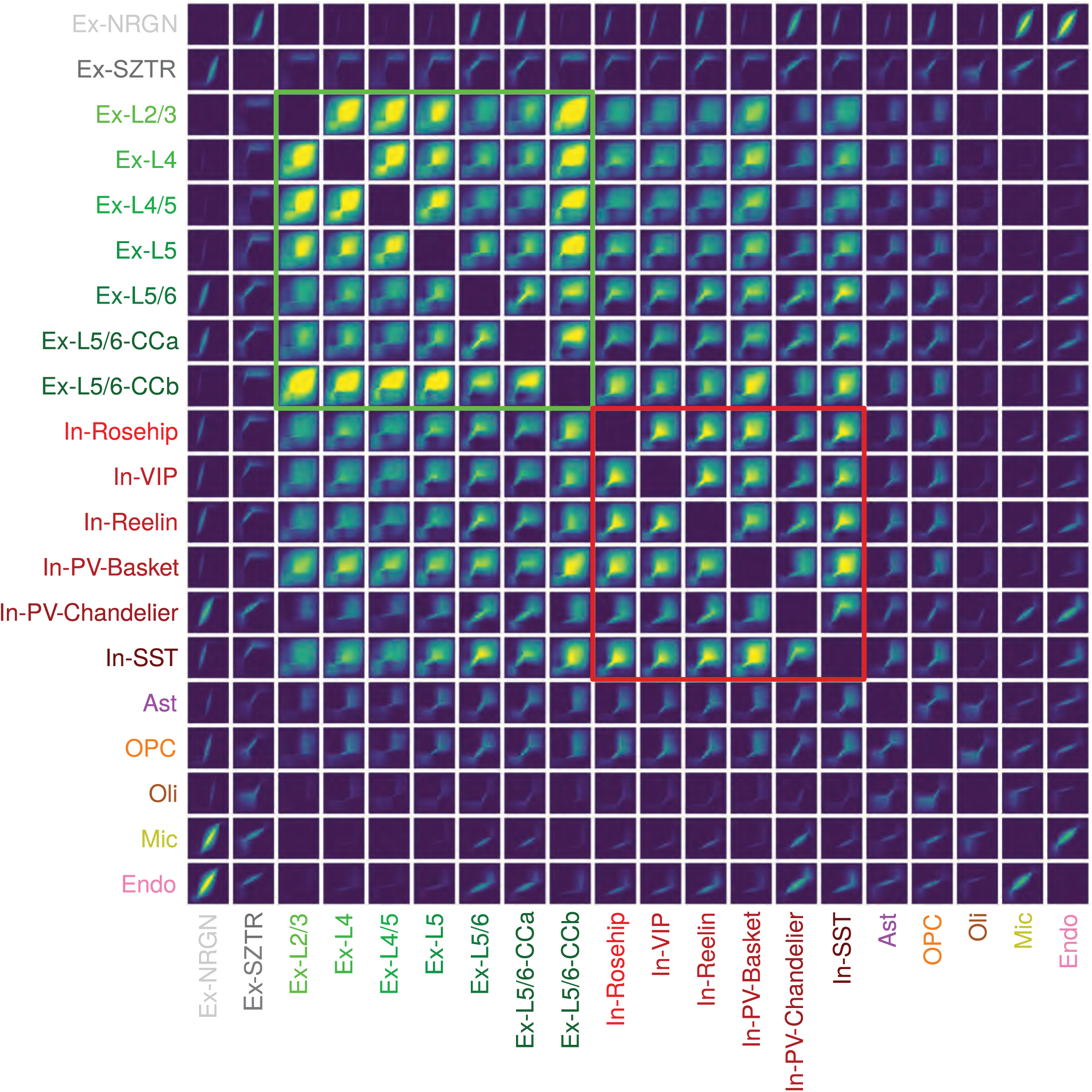
Rank-Rank Hypergeometric Overlap Plot of cell-type-specific trancriptomic changes. Rank-Rank Hypergeometric Overlap plot depicting the similarity of transcriptional perturbations between all pairs of cell-types/states.

**Extended Data Figure 4.**
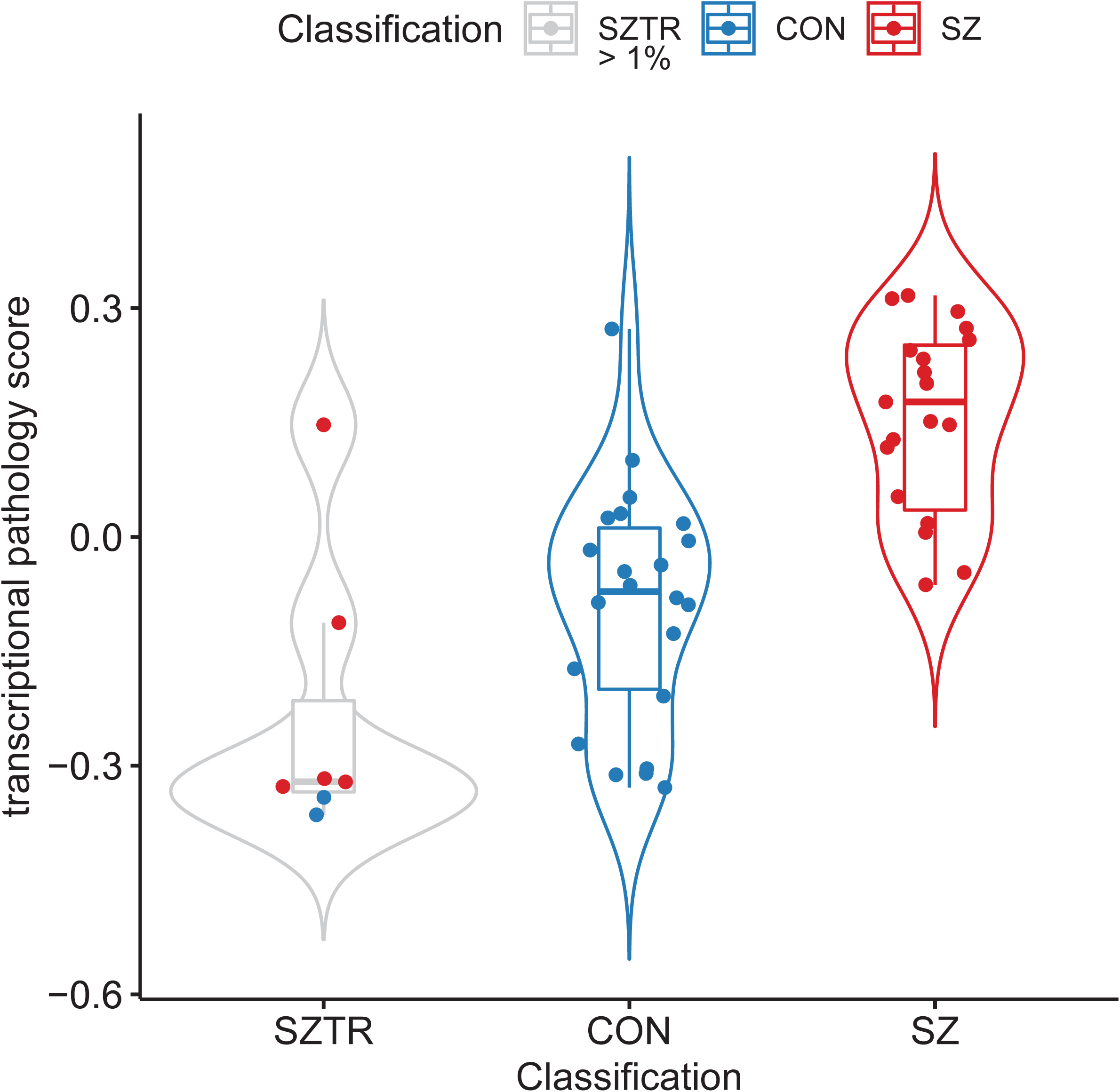
Transcriptional pathology scores across subject classification. Transcriptional pathology scores across all individuals demonstrate clear gradation across classifications, with SZTR individuals (Ex-SZTR cell fraction > 1%) ranking below the majority of CON individuals, away from the SZ group.

**Extended Data Figure 5.**
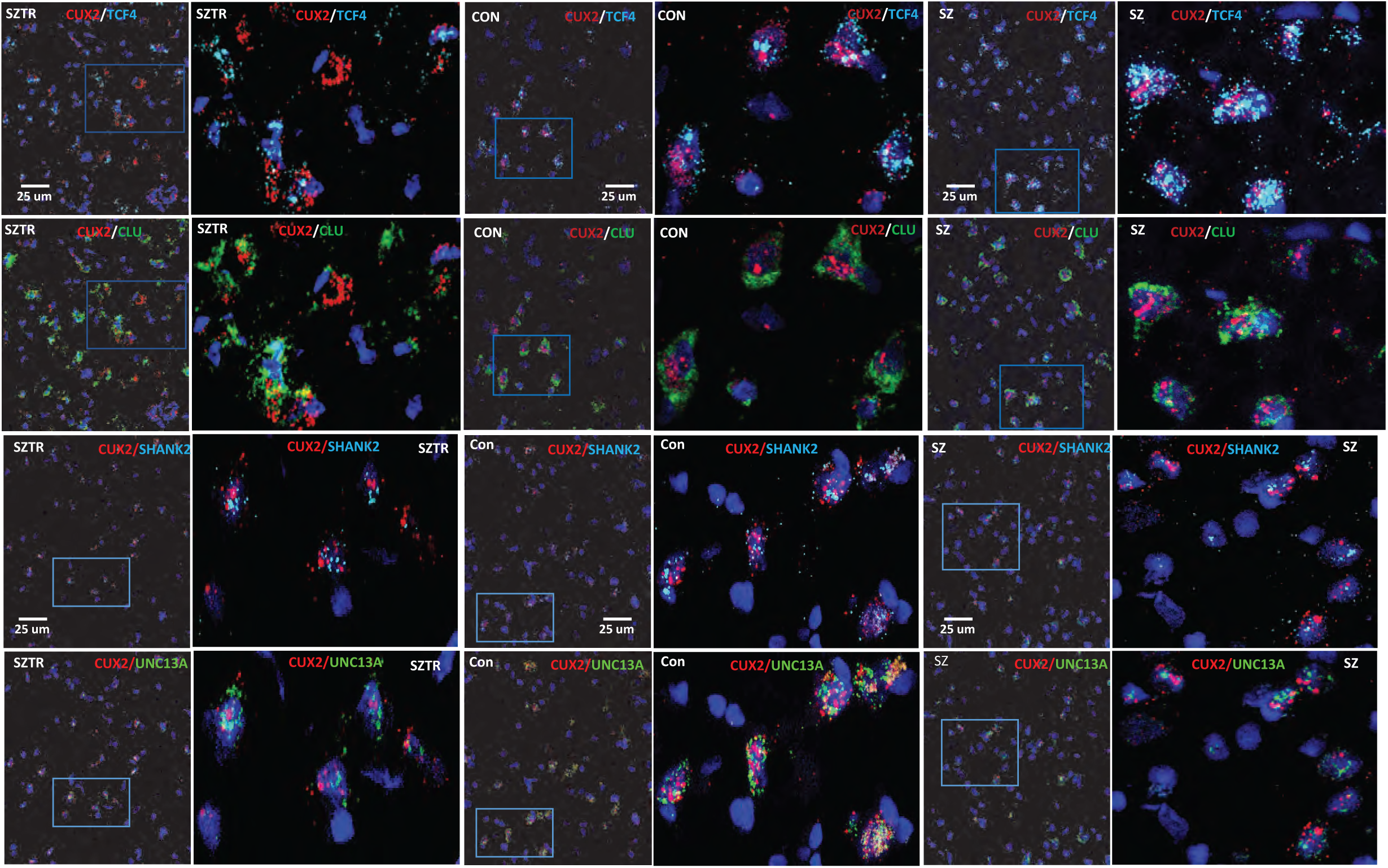
Fluorescence *in situ* hybridization of selected SZ DEGs. Representative photomicrographs from one case in each phenotype (SZTR, CON, SZ) are shown of in situ hybridization for the *CUX2* (Red, layer II and III excitatory neuron marker), *TCF4* (top, blue), *CLU* (middle, green), *SHANK2* (middle, blue), and *UNC13A* genes (bottom, green). Each image was captured at 40x magnification and for each image the full field of view is shown to the left, and the area boxed in blue is shown to the right. Each gene was assessed in two cases from each phenotype for six total cases.

**Extended Data Figure 6a.**
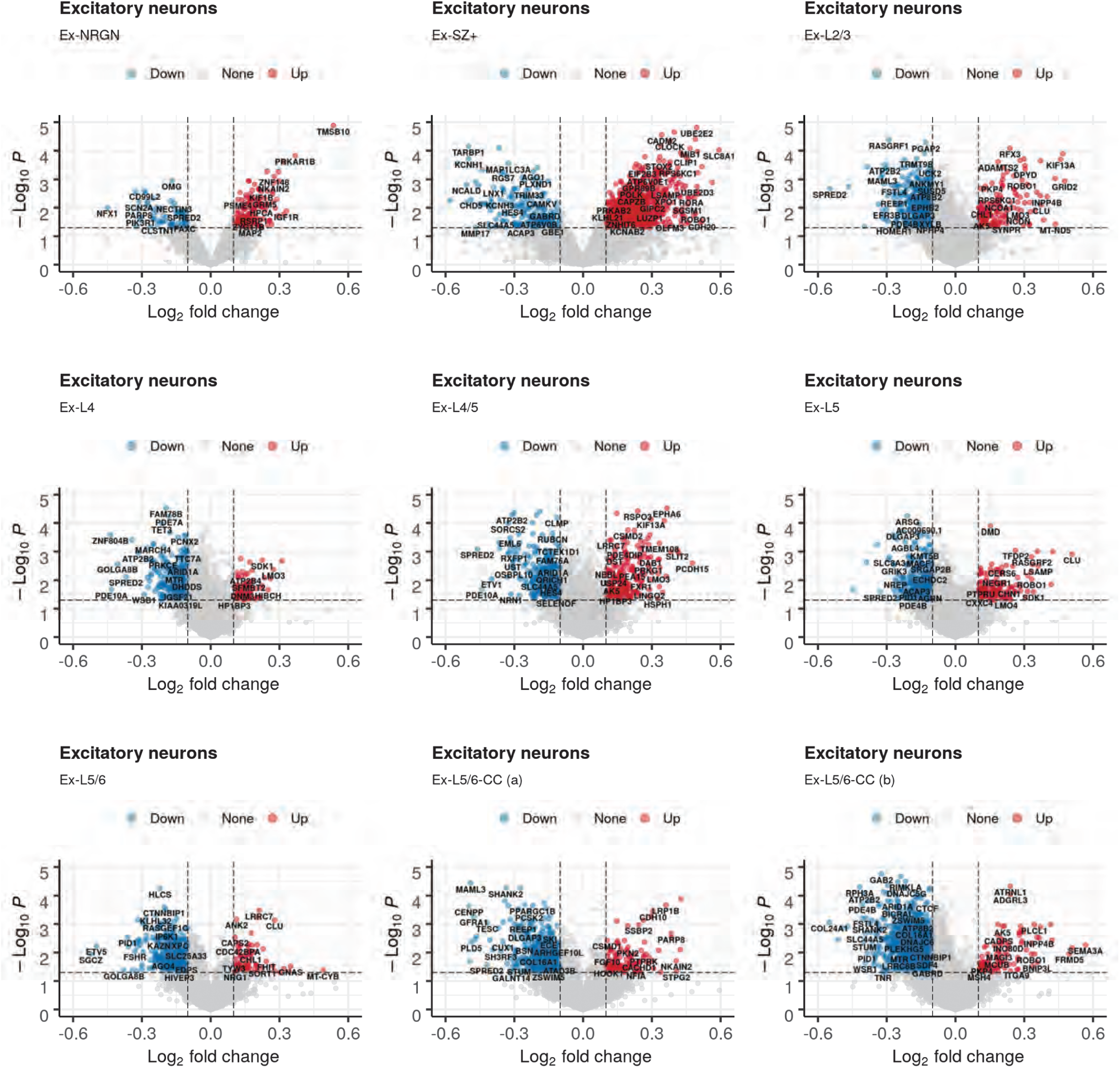
Volcano plots of SZ GWAS significance versus cell-type-specific transcriptional perturbation in excitatory neuronal populations. **Genetic versus transcriptional SZ-associated perturbations**. Volcano plots showing the relationship between SZ GWAS scores^18^ (y-axis, −log10 association p-value) and SZ transcriptional perturbations measured and reported herein (x-axis, Log2 fold change of expression values in SZ relative to control samples). Plots are shown independently for subpopulations of excitatory neurons.

**Extended Data Figure 6b.**
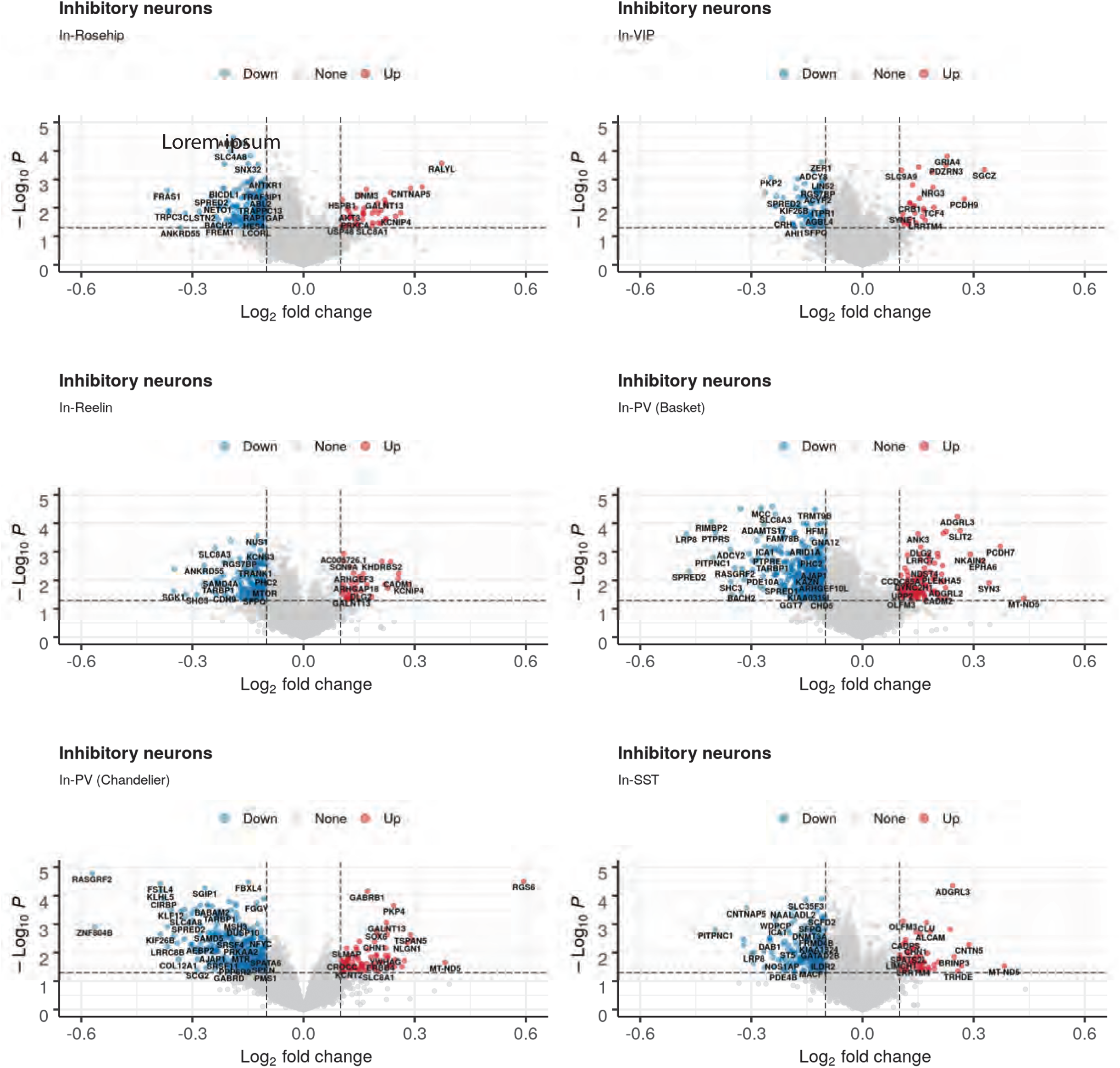
Volcano plots of SZ GWAS significance versus cell-type-specific transcriptional perturbation in inhibitory neuronal populations. **Genetic versus transcriptional SZ-associated perturbations**. Volcano plots showing the relationship between SZ GWAS scores^18^ (y-axis, −log10 association p-value) and SZ transcriptional perturbations measured and reported herein (x-axis, Log2 fold change of expression values in SZ relative to control samples). Plots are shown independently for subpopulations of inhibitory neuron subpopulations.

**Extended Data Figure 6c.**
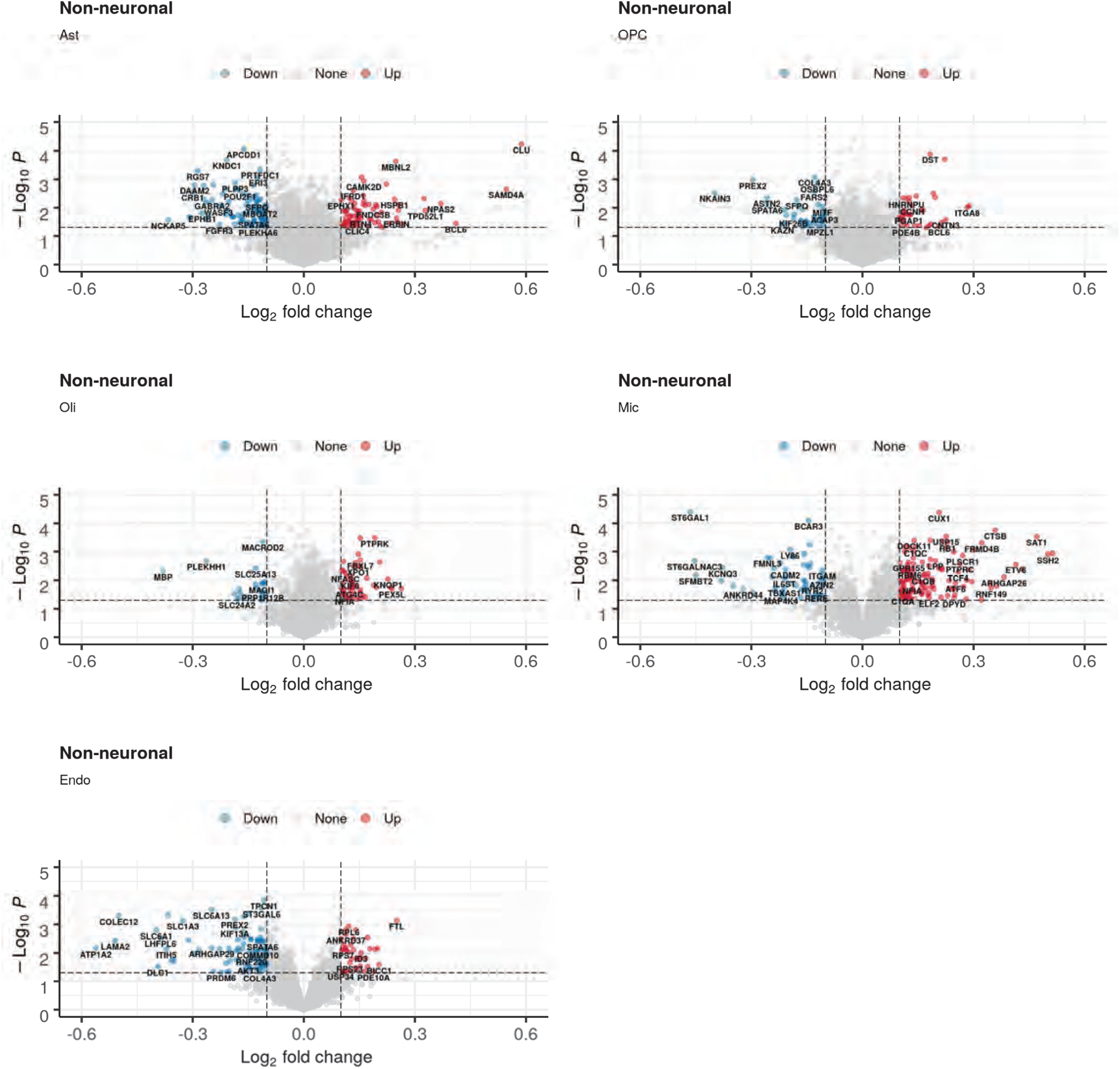
Volcano plots of SZ GWAS significance versus cell-type-specific transcriptional perturbation in non-neuronal populations. **Genetic versus transcriptional SZ-associated perturbations**. Volcano plots showing the relationship between SZ GWAS scores18 (y-axis, −log10 association p-value) and SZ transcriptional perturbations measured and reported herein (x-axis, Log2 fold change of expression values in SZ relative to control samples). Plots are shown independently for subpopulations of non-neuronal subpopulations.

**Extended Data Figure 7.**
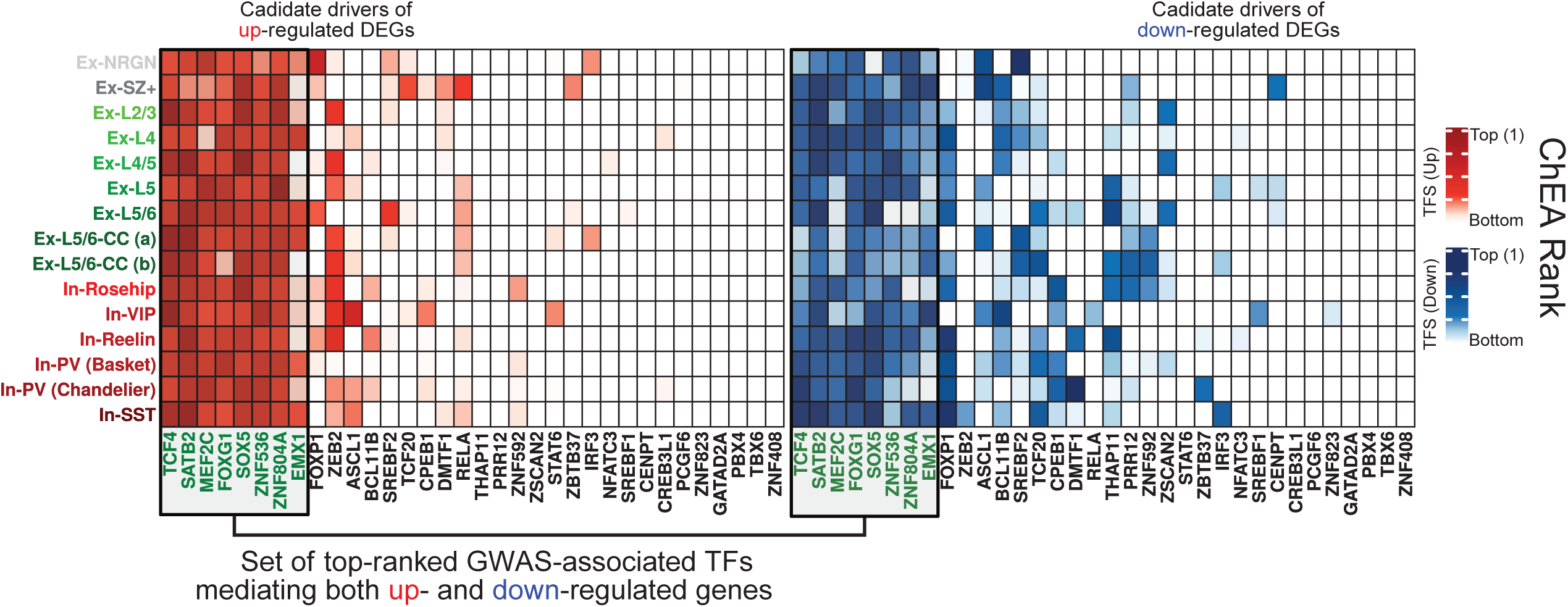
Enrichment of SZ GWAS TF targets within SZ DEGs. Overrepresentation analysis (hypergeometric test) within targets of TFs genetically associated with SZ in GWAS (columns) of genes detected as differentially expressed in SZ relative to controls (rows). Overrepresentation analysis was performed independently for SZ upregulated (left) and downregulated genes (right).

**Extended Data Figure 8.**
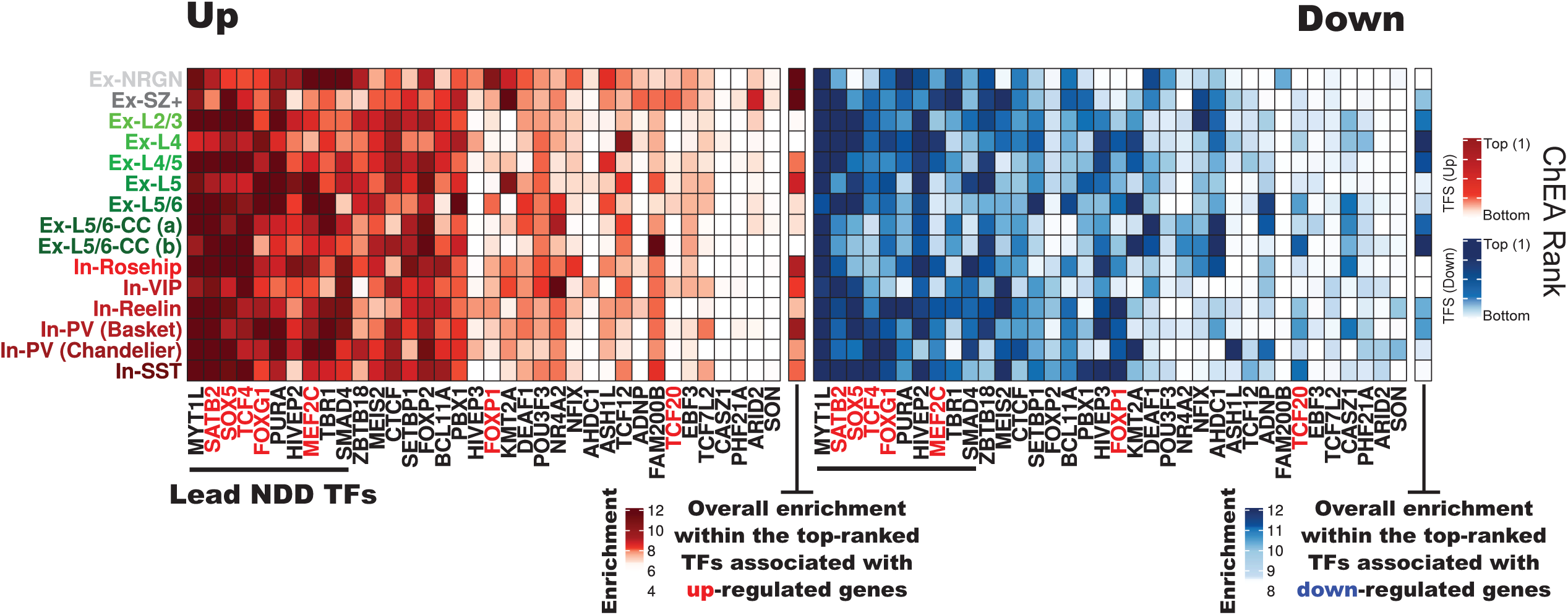
Enrichment of neurodevelopmental delay-associated TF targets within SZ DEGs. Overrepresentation analysis (hypergeometric test) within targets of TFs genetically associated with neurodevelopmental delay65 (de novo mutations and CNVs) (columns) of genes detected as differentially expressed in SZ relative to controls (rows). Overrepresentation analysis was performed independently for SZ upregulated (left) and downregulated genes (right).

**Extended Data Figure 9.**
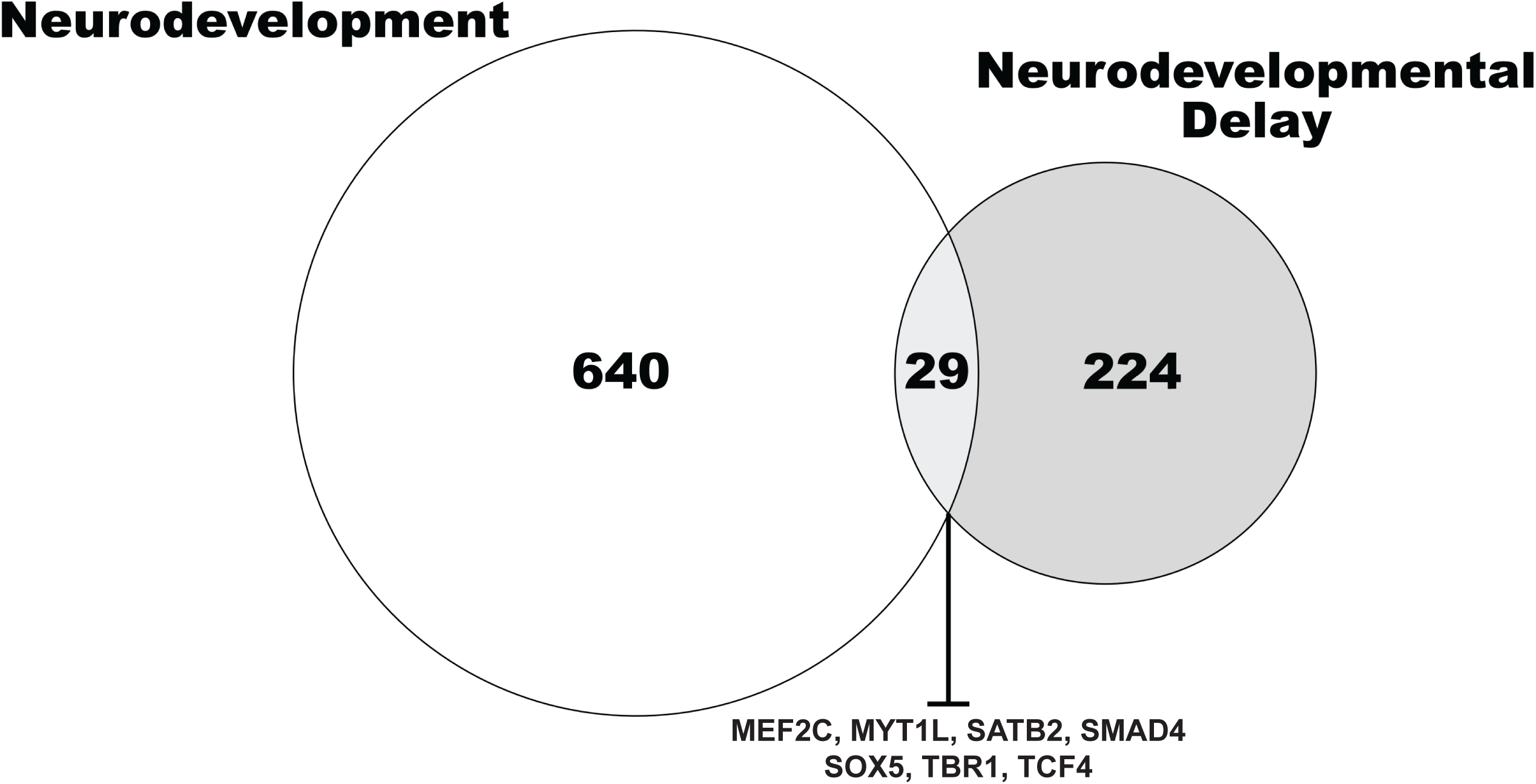
Neurodevelopment and neurodevelopmental delay associated genes overlap. Overlap of genes genetically associated with neurodevelopmental delay (NDD) and genes functionally annotated as related with neurodevelopment. SZ TFs are associated with both genesets. The majority of the lead NDD TFs (7/10) are also involved in neurodevelopment.

**Extended Data Figure 10.**
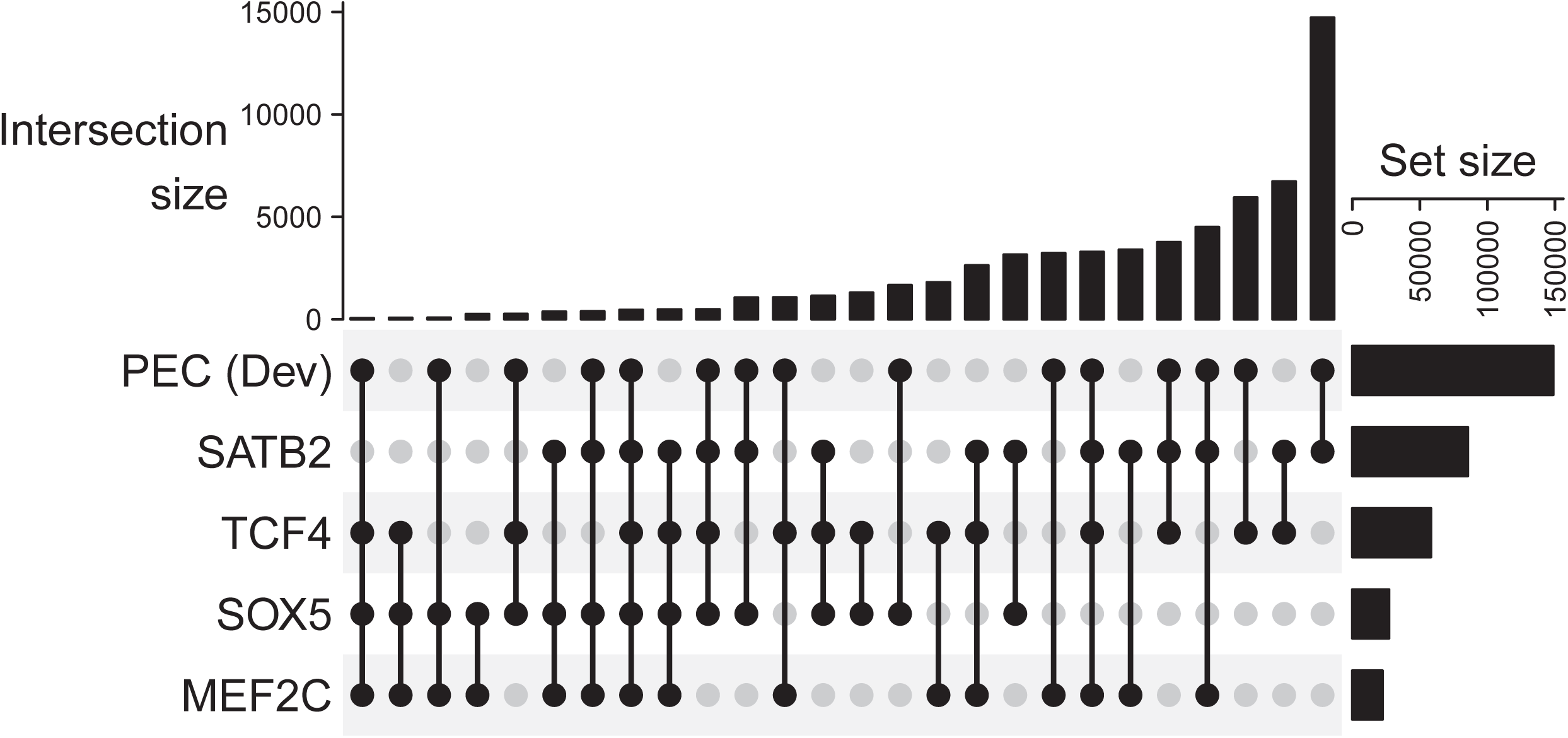
Overlap of PEC developing human brain regulatory elements and observed TF binding sites in neuronal nuclei. The overlap of the total number of peaks for each transcription factor, defined as the union of top 1% peaks observed in any of the samples, with the masterset of PEC developing human brain dataset, containing H3K27Ac peaks for human brain samples at different stages of development.

